# Effects of polygenic risk for suicide attempt and risky behavior on brain structure in young people with familial risk of bipolar disorder

**DOI:** 10.1101/2021.09.06.21262817

**Authors:** Bronwyn J. Overs, Gloria Roberts, Kate Ridgway, Claudio Toma, Dusan Hadzi-Pavlovic, Holly C. Wilcox, Leslie A. Hulvershorn, John I. Nurnberger, Peter R. Schofield, Philip B. Mitchell, Janice M. Fullerton

**Affiliations:** Neuroscience Research Australia, Randwick, New South Wales, Australia; School of Psychiatry, University of New South Wales, Kensington, New South Wales, Australia; Centro de Biología Molecular “Severo Ochoa”, Universidad Autónoma de Madrid/CSIC, Madrid, Spain; Child Psychiatry & Public Health, Johns Hopkins University, Baltimore, Maryland; Department of Psychiatry, Institute of Psychiatric Research, Indiana University School of Medicine, Indiana, Indianapolis; Department of Medical and Molecular Genetics, Indiana University, Indiana, Indianapolis; School of Medical Sciences, University of New South Wales, New South Wales, Kensington, Australia

**Author notes:** Corresponding Author: Janice M. Fullerton, Neuroscience Research Australia, Margarete Ainsworth Building, Barker Street, Randwick, Sydney, NSW 2031, Australia; E. Grant Numbers: Australian National Medical and Health Research Council (NHMRC) Program Grant 1037196, Project Grant 1066177, Investigator Grants 1176716 and 1177991.

**Keywords:** polygenic risk score, structural magnetic resonance imaging, cuneus, parahippocampus, anterior cingulate.

## Abstract

**Aims:** Bipolar Disorder (BD) is associated with a 20-30 fold increased suicide risk compared to the general population. First-degree relatives of BD patients show inflated rates of psychopathology including suicidal behaviors. As reliable biomarkers of suicide attempts (SA) are lacking, we examined associations between suicide-related polygenic risk scores (PRS) – a quantitative index of genomic risk – and variability in brain structures implicated in SA.

**Methods:** Participants (n=206; aged 12-30 years) were unrelated individuals of European ancestry and comprised three groups: 41 BD cases, 96 BD relatives (‘high-risk’), and 69 controls. Genotyping employed PsychArray, followed by imputation. Three PRS were computed using genome-wide association data for SA in BD (SA-in-BD), SA in Major Depressive Disorder (SA-in-MDD) [Mullins et al., 2019], and risky behavior [Karlsson Linnér et al., 2019]. Structural MRI processing employed FreeSurfer v5.3.0. General linear models were constructed using 32 regions-of-interest identified from suicide neuroimaging literature, with false-discovery-rate correction.

**Results:** SA-in-MDD and SA-in-BD PRS negatively predicted parahippocampal thickness, with the latter association modified by group membership. SA-in-BD and Risky Behavior PRS inversely predicted rostral and caudal anterior cingulate structure, respectively, with the latter effect driven by the ‘high-risk’ group. SA-in-MDD and SA-in-BD PRS positively predicted cuneus structure, irrespective of group.

**Conclusions:** This study demonstrated associations between PRS for suicide-related phenotypes and structural variability in brain regions implicated in SA. Future exploration of extended PRS, in conjunction with a range of biological, phenotypic, environmental and experiential data in high-risk populations, may inform predictive models for suicidal behaviors.

## Introduction

Bipolar disorder (BD) is a highly heritable mood disorder with a lifetime prevalence of ∼1-4% (Kessler et al., 2006; Merikangas et al., 2007; Merikangas et al., 2011), characterised by episodes of elevated and low mood (mania and depression), increased impulsivity and risk-taking behavior (Ramirez-Martin, Ramos-Martin, Mayoral-Cleries, Moreno-Kustner, & Guzman-Parra, 2020). BD is also associated with a reduced life expectancy of 8 to 10 years compared to the general population (Kessing, Vradi, & Andersen, 2015). Suicide accounts for a substantial proportion of premature mortality in BD, with 20-60% of BD patients attempting suicide at least once within their lifetime, and 5-20% of BD mortality being attributed to suicide (Goodwin & Jamison, 2007; Ilgen et al., 2010; Rihmer, Gonda, & Döme, 2017). In addition, risk of suicide-related mortality in BD is between 20 to 30 times higher than suicide-related mortality in the general population (Plans et al., 2019), suggesting stronger suicide intent and greater lethality of chosen methods. Furthermore, first-degree relatives of BD patients carry a 5-10 fold increased risk of developing BD themselves (Craddock & Sklar, 2013; Smoller & Finn, 2003) and are generally identified as being at increased risk of broader psychopathology. Significantly, such ‘high-risk’ individuals exhibit substantially increased rates of suicidal ideation (SI) and suicide attempts (SA) relative to the general population, even after adjusting for personal mood and substance use disorders (Wilcox et al., 2017); these findings suggest that genetic factors contribute to suicidal behaviors independently of mood disorder symptoms.

In recent years, efforts to establish the genetic architecture underlying suicide behaviors have made substantial progress. Early twin studies have indicated that the heritability of suicide is approximately 30-55% (Voracek & Loibl, 2007), with additive genetic predictors that are largely distinct from those implicated in the aetiology of any underlying psychiatric disorder (Brent & Mann, 2005). When heritability for other psychiatric conditions was accounted for, the residual heritability estimate was 17.4% (Fu et al., 2002); indicating that suicide-specific genetic factors exist. Family studies have shown that the risk of suicide attempts is higher in relatives of people who died by suicide, and that the risk of dying by suicide is higher in relatives of people with a history of suicide attempts (Turecki & Brent, 2016). Several gene discovery studies have been conducted, identifying common variants (Erlangsen et al., 2020; Galfalvy et al., 2015; Mullins et al., 2019; Mullins et al., 2014; Otsuka et al., 2019; Perlis et al., 2010; Ruderfer et al., 2020; Schosser et al., 2011; Stein et al., 2017; Strawbridge et al., 2019; Willour et al., 2012) and specific rare variants (Coon et al., 2020; DiBlasi et al., 2021) associated with SA or suicide-related mortality. The latest genome-wide association study (GWAS) of suicide behavior from the Psychiatric Genomics Consortium (PGC) – involving 6,569 ‘case’ participants (with diagnoses of major depressive disorder [MDD], BD or schizophrenia, who reported suicide attempts), compared to 17,232 ‘control’ participants (with the same diagnoses but who had no history of suicidal behaviour) – identified one locus on chromosome 10 for SA in MDD (MDD attempters n = 1,622; MDD non-attempters n = 8,786) and one locus on chromosome 4 for SA in BD (BD attempters n = 3,264, BD non-attempters n = 5,500), that met genome-wide significance (Mullins et al., 2019). In addition, meta-analysis of SA across mood disorders (MDD and BD) revealed two loci that met genome-wide significance on chromosomes 2 and 4 (Mullins et al., 2019). Several genetic studies have also highlighted the polygenic nature of genetic determinants of SA. Specifically, polygenic risk scores (PRS) derived from discovery GWAS leave-one-out summary statistics demonstrated validity of genomic signals (Mullins et al., 2019), and PRS derived from other GWAS for SA or suicide death (Docherty et al., 2020) have shown a moderate ability to predict SA within independent cohorts (Erlangsen et al., 2020; Mullins et al., 2014; Ruderfer et al., 2020). While these findings provide tentative evidence for the existence of genetic biomarkers for SA, derived PRS account for less than 1% of variability in the logistic model odds ratios of SA using pseudo-R^2^, thus further work is required to establish a robust biological index that may be utilized within a clinical setting to reliably predict suicide behaviors.

Examination of the genetic determinants of clinical features that are strongly associated with SA via the ideation-to-action pathway may also prove critical to identifying which individuals are most vulnerable to SA. Given the established association between higher engagement in risky behaviors and SA (Eaton et al., 2011; Pena, Matthieu, Zayas, Masyn, & Caine, 2012; Thullen, Taliaferro, & Muehlenkamp, 2016), genetic determinants of behavioral disinhibition, impulsivity and risky behavior (including alcohol abuse, illicit drug use, risky sexual behavior and interpersonal violence) warrant further investigation in relation to biomarkers for suicide risk. A recent systematic review found significant impairment of various aspects of impulsivity in BD, many of which persist beyond mood symptoms, suggesting that impulsivity, decision-making and risk-taking may represent core clinical features that are highly relevant to the strong links between BD and suicidal mortality (Ramirez-Martin et al., 2020). In addition, risky behaviors have also been more readily observed in both BD (Gordon-Smith et al., 2020; Hunt, Malhi, Cleary, Lai, & Sitharthan, 2016; Obo, Sori, Abegaz, & Molla, 2019; Volavka, 2013) and familial risk for BD (Nijjar, Ellenbogen, & Hodgins, 2014), and may account for some of the increased incidence and lethality of SA in BD. While estimated heritability of risk tolerance (i.e. willingness to take risks, usually in receipt of a reward) has varied between twin studies, estimates of relevant risk behavior factors have ranged from .30 to .66 (Beauchamp, Cesarini, & Johannesson, 2017; Harden et al., 2017). To date, the largest GWAS of general risk tolerance and risky behavior involved a combined sample of 939,908 participants from the UK Biobank and 23andMe (Karlsson Linnér et al., 2019). This study identified 99 genome-wide significant loci associated with general risk tolerance, and a PRS derived from GWAS discovery and replication samples accounted for 1.6% of the variation in general risk tolerance using pseudo-R^2^. The availability of this large GWAS of risky behavior facilitates the novel exploration of relationships between additive polygenic profiles for risky behavior and neuroimaging biomarkers for SA.

Over recent decades, a large body of research has focused on the identification of neuroimaging biomarkers that precede or are associated with suicide-related behaviors (Schmaal et al., 2020). Cortical and subcortical findings related to suicidal thoughts and behaviors appear to converge on reduced volume and thinner cortex in brain systems that involve impulse and emotion regulation, including the ventral and dorsal prefrontal cortex, insula and anterior cingulate, as well as their primary and secondary connection sites; these findings may be important to the blunted positive and excessive negative internal states that can stimulate SI (Schmaal et al., 2020). Furthermore, there is strong evidence of cross-diagnostic features of suicidality impacting the default mode, affective and reward networks, with consistency of findings across the psychiatric disorders (Malhi et al., 2020; Minzenberg et al., 2015), indicating that the mechanisms underlying suicidal behaviors may be independent of other clinical symptoms.

Thus far, clinical tools for suicide prediction have rarely examined the combined effect of multiple risk factors (Franklin et al., 2017), have relied mostly on demographic information, family history and clinical symptoms, and have shown poor predictive value (Kessler, Bossarte, Luedtke, Zaslavsky, & Zubizarreta, 2020), suggesting the need for novel multimodal approaches for improved risk prediction. Furthermore, distinct mechanistic processes are posited to underlie SI and SA (Wiebenga, Eikelenboom, Heering, van Oppen, & Penninx, 2021), with differing environmental, social, physiological and biological factors – such as sensitivity to pain, impulsivity, control and conflict awareness – that influence experiences leading up to and during suicidal crises (Klonsky & May, 2014); these processes may moderate the ideation-to-action pathway. While research has supported the role of various biological factors in the etiology of suicidal behaviors (Sudol & Mann, 2017), examination of the relationships between different types of potential biomarkers is lacking. Further, it has been suggested that unimodal approaches, which attempt to predict suicidality using a single type of biomarker (e.g. genetic or neuroimaging alone), cannot sufficiently represent the complex biological profiles that are associated with suicidal behaviors (Schmaal et al., 2020). Therefore, the examination of relationships between discrete biological factors that precede SA may illuminate the ideation-to-action pathway, facilitating the development of reliable clinical tools, with sufficiently high sensitivity and specificity, that may assist in reducing suicide mortality. The present study sought to undertake the novel examination of relationships between potential genetic and neuroimaging biomarkers of SA. We aimed to establish if PRS derived from GWAS of SA (Mullins et al., 2019) and risky behavior (Karlsson Linnér et al., 2019) could predict structural variability in brain regions-of-interest (ROI) that were defined by previous neuroimaging literature in SA. This analysis was performed within a European ancestry sample of 206 unrelated participants, including BD patients, young first-degree relatives of BD patients (who were not confounded by a personal diagnosis of BD and were not related to the BD patients in the present study), and control participants with no personal or family history of BD; this facilitated the examination of interactions between clinical group and polygenic risk, in the prediction of brain ROI structure.

## Methods

### Participants

All participants were aged between 12 and 30 years and were part of an ongoing longitudinal study of familial risk of BD, as detailed elsewhere (Perich et al., 2015). Exclusion criteria included missing data across clinical, MRI or genotype modalities (n=43), genotype quality control failure (n=3; as detailed below), MRI quality control failure (n=12; as detailed below), familial relation to an included participant (n=49), and non-European ancestry as determined by self-report and multi-dimensional scaling analysis of genomic data (n=74; as detailed below). The final analysis sample comprised 206 unrelated participants in three groups: 1) 41 BD participants with an established diagnosis of either BD-I or BD-II; 2) 96 ‘high-risk’ individuals, who had a first degree relative with BD-I, BD-II or schizoaffective disorder (bipolar-type), but had no personal history of these three psychiatric disorders; and 3) 69 control participants who were defined as those who did not have a first-degree relative with either BD-I or II, schizophrenia, schizoaffective disorder, recurrent major depression, recurrent substance abuse, psychosis or psychiatric hospitalization. Additionally, controls did not have a second-degree relative with a history of psychosis or hospitalization for a mood disorder. ‘High-risk’ and control participants with a lifetime or current presence of psychiatric symptoms (apart from the occurrence of BD) were not excluded from the study. Ethical approval to conduct this study was provided by the University of New South Wales Human Research Ethics Committee (HREC Protocol 09/097). Written informed consent was obtained from all participants with additional parental consent for participants aged 16 or younger.

### Clinical and Cognitive Measures

To determine family history of BD and other psychiatric disorders, the Family Interview for Genetic Studies (FIGS) (Maxwell, 1992) was administered to each participant or a member of their family. Parents of 12 to 21 year old participants were interviewed about their child using the Kiddie-Schedule for Affective Disorders and Schizophrenia for School-Aged Children – Present and Lifetime Version (K-SADS-BP; Version 2, 24 July 2009)(Nurnberger et al., 2011), in addition to the participant themselves completing a K-SADS-BP interview. For all participants aged 22 to 30 years and BD-probands of ‘high-risk’ participants, the Diagnostic Interview for Genetic Studies (DIGS; Version 4.0/BP, 21 July 2005) was administered (Nurnberger et al., 1994). Consensus DSM-IV diagnoses for BD, ‘high-risk’, BD-probands of ‘high-risk’ and control participants were determined by two independent raters, using best estimate methodology (Leckman, Sholomskas, Thompson, Belanger, & Weissman, 1982), drawing from the DIGS, K-SADS-BP, FIGS and medical records (where available). The presence of any lifetime SI was determined by item 26 of the KSADS-BP section on depression or items 15 and 48 of the DIGS sections on most severe and other major depression episodes (Section F). The presence of any lifetime SA was determined by item 27 of the KSADS-BP section on depression or items 16 and 49 of the DIGS sections on most severe and other major depression episodes (Section F), as well as a later section of the DIGS on suicidal behavior more broadly (items 1-18 in Section O). The Children’s Depression Inventory (CDI) (Kovacs, 1992) and the Montgomery– Åsberg Depression Rating Scale (MADRS) (Montgomery & Asberg, 1979) were used to assess current mood state at baseline for participants aged 21 years and below, and 22 years and above (respectively). The Young Mania Rating Scale (YMRS) (Young, Biggs, Ziegler, & Meyer, 1978) was used to examine baseline manic symptoms in participants aged 22 years and above; baseline mania was not examined in participants aged 21 years and below. Finally, the Wechsler Abbreviated Scale of Intelligence (WASI) (Wechsler, 1999) was used to assess intellectual quotient (IQ) at baseline for all participants.

### Genotyping and Polygenic Risk Score Calculation

Genomic DNA was extracted from peripheral blood samples by standard procedures, and genome-wide SNP genotyping conducted using the Infinium PsychArray BeadChip v1 (Illumina, Scoresby, Victoria, Australia), as previously described (Jamshidi et al., 2020; Wilcox et al., 2017). PsychArray genotype calling and quality control were conducted in two batches using standard PGC Ricopili pipeline (v8.0), incorporating consensus-merge genotype calls from three algorithms (birdseed, zCall, genCall), as previously described (Overs et al., 2021; Wilcox et al., 2017). In brief, SNPs with Hardy-Weinberg Equilibrium (HWE) p<1×10^-06^ or genotype call rates < 97% were removed, before merging resultant files in PLINK v1.9 (Purcell et al., 2007). A second quality control was then applied to remove individuals with inbreeding coefficient outside ±.2 or genotype-derived sex discrepancy with clinical files, and individuals with SNP call rates <98%. SNPs with HWE p<1×10^-6^ in controls or HWE p<1×10^-10^ in cases and minor allele frequency (MAF) <.00001 were excluded, leaving 426,091 SNPs. The successfully genotyped SNPs had a >99.6% genotype pass rate.

Genotype-derived ancestry was assessed by multi-dimensional scaling (MDS) analysis of genotype data, implemented in PLINK v1.9 (Purcell et al., 2007), following ENIGMA2 protocols (http://enigma.ini.usc.edu/protocols/genetics-protocols/ [13 July2017]). Briefly, post-QC genotype files were merged with genotypes from ethnically diverse 1000 Genomes samples (Human Mapping Set, phase 3 release v5). MDS analysis employed 164,680 independent SNPs, from which four principal components were calculated. The values of the first two principal components from each participant were compared to those of subjects of known ancestry, to establish a genotype-derived ancestry for each participant (Figure S1). Identity-by-descent estimates were generated from 75,182 independent SNPs and were used to confirm that no spurious familial relationships existed between subjects (pi_hat<0.125, excludes up to 3^rd^ degree relatives).

Prior to haplotype phasing and imputation using the Michigan Imputation Server (v1.0.3) (Das et al., 2016) (https://imputationserver.sph.umich.edu/), SNPs with MAF <.01, genotype call rate <95%, strand ambiguity and those with HWE p<1e^-06^ were excluded. The 1000 Genomes Phase 3 v5 reference panel (European population) was employed, with phasing by Eagle v2.3 in quality control and imputation mode. The reference overlap of uploaded SNPs was 99.65%, and the allele frequency correlation of reference panel against uploaded samples was R^2^ =.993. Post-imputation files contained approximately 47 million variants. To retain common, high quality imputed genotypes for downstream analysis, SNPs with imputation quality score Rsq <.8, MAF <.05 and HWE p<1×10^-6^ were removed. After imputation, files were merged with imputed genotype outputs from the Genetic Association Information Network (GAIN) BD sample (http://zork.wustl.edu/nimh/)(Smith et al., 2009) and BOMA-Australia cohorts (Muhleisen et al., 2014), which included parental relatives of ‘high-risk’ offspring, yielding a total of 9,238 participants. Risk scores were generated across the whole cohort (for consistency of reporting), but the current study employed analysis of a subset of those unrelated Australian participants whose neuroimaging data were available. Imputed names for 4,028,658 SNPs were recoded from hg19 “chr:positions” to an rsID using the file snpinfo_1kg_hm3 (included within the ldblk_1kg_eur.tar.gz package; available from https://www.dropbox.com/s/mt6var0z96vb6fv/ldblk_1kg_eur.tar.gz).

Polygenic risk scores (PRS) were generated using all SNPs (i.e. p-value threshold of 1), employing the PRS-CS algorithm (https://github.com/getian107/PRScs; version 10 Sept, 2020), a Python-based tool that infers posterior SNP effect sizes under continuous shrinkage priors, using GWAS summary statistics and an external reference panel (1000 Genomes EUR; ldblk_1kg_eur.tar.gz, downloaded 30 Oct, 2020) to enable multivariate modelling of local linkage disequilibrium patterns in the generation of a genomic index of allelic risk for each participant (Ge, Chen, Ni, Feng, & Smoller, 2019). The latest version of Python was employed (anaconda3-2020.07-Linux-x86_64; downloaded 13 Nov 2020).

### GWAS summary statistics were obtained for

- SA-in-MDD – (Mullins et al., 2019) Discovery GWAS comprised 1,622 suicide attempters and 8,786 non-attempters with MDD, and sumstats were downloaded from PGC website (https://www.med.unc.edu/pgc/download-results/).
- SA-in-BD – (Mullins et al., 2019) Discovery GWAS comprised 2,911 attempters and 5,099 non-attempters with BD, after excluding FAT2 and GAIN cohorts, which contained parents of some of the US-based ‘high-risk’ study participants. Leave-one-out sumstats were received via personal communication (Niamh Mullins, on 11 Aug 2020).
- Risky Behavior – (Karlsson Linnér et al., 2019) Discovery GWAS for general-risk-tolerance were derived from a GWAS meta-analysis of the UK Biobank and 10 replication cohorts (excluding the 23andMe cohort) comprising 466,571 participants, and sumstats were downloaded from (https://www.thessgac.org/data).

All sumstats files were pruned to exclude strand ambiguous SNPs (and where available, INFO<.6 and MAF<.05, yielding ∼815,000 SNPs), and effect allele was coded as A1. PRS-CS was run using default parameters (fixing phi to 1e for polygenic traits). Posterior SNP effect size estimates generated by PRS-CS were used as input to PLINK, with --score function to generate individual-level PRS for each participant. For each PRS, scores were derived from an average of ∼417,000 alleles, and were standardized (within the neuroimaging sample, n=206) to a mean of 0 and standard deviation of 1.

### Structural Imaging Acquisition and Processing

T1-weighted structural MRIs were acquired for each participant at baseline using a 3T Philips Achieva scanner (Royal Philips Electronics, Amsterdam, the Netherlands). Each MRI comprised 180 sagittal T1-weighted three-dimensional turbo field-echo images (TR/TE – 5.5/2.5ms, flip angle – 8°, field of view – 256×256×180mm^3^ , voxel size – 1×1×1mm, scan time – 371s). To account for head positioning and resolve sensitivity variations, a one-minute standard scout image was taken prior to image acquisition.

Prior to sample finalisation, all MRI images were subjected to a multi-step quality control (QC) procedure that began with manual image inspection for brain abnormalities and artefacts (motion, contrast, and orientation). For images that passed manual inspection, automated cortical reconstruction and segmentation was undertaken using Freesurfer version 5.3.0 (http://surfer.nmr.mgh.harvard.edu/); this involved Talairach registration, intensity normalisation, skull stripping, surface tessellation, cortical parcellation, subcortical segmentation, and topology correction (Dale, Fischl, & Sereno, 1999; Desikan et al., 2006; Fischl, Liu, & Dale, 2001; Fischl et al., 2002). Post processing QC of reconstructed images was completed using standardized procedures for the Freesurfer software package. A threshold of four or more erroneous voxels across two slices was employed for editing of pial and white matter boundaries to be undertaken. For all imaging QC, procedures were undertaken by three research team members who were blinded to participant group and demonstrated inter-rater reliability of greater than .9 on average (for all Desikan-Killiany parcellated volumes, across a set of twenty participant MRIs).

For images that passed the above QC procedures, measures of cortical area, thickness and volume were extracted for all 68 regions (34 regions per hemisphere) defined by the Desikan-Killiany atlas (Desikan et al., 2006; Fischl et al., 2004), as well as measures of volume for all subcortical regions segmented during the Freesurfer processing stream (22 regions, 11 per hemisphere, excluding the ventricles, brain stem, cerebrospinal fluid and vessel). For 7 participants (3 control, 4 BD) who demonstrated persistent pial boundary errors that exceeded 100 voxels in size (during QC), measures of thickness, area, and volume for the affected cortical regions (left caudal anterior cingulate, rostral anterior cingulate, isthmus of the cingulate, and/or posterior cingulate, and/or right medial orbitofrontal) were replaced with missing values, excluding these participants from subsequent analysis of these specific regions.

To determine target regions-of-interest (ROIs) that have previously shown structural imaging differences following SA, a literature review was conducted in December 2020 (Table 1). Building on the work of Cox Lippard, Johnston and Blumberg (2014), we employed PubMed (https://pubmed.ncbi.nlm.nih.gov/) using search terms including ‘Bipolar’, ‘Suicide’, ‘Suicide Attempt’, ‘MRI’, ‘Magnetic Resonance Imaging’, ‘Brain’ and ‘Structural’, used in varying combinations. Search results were manually examined and publications were retained for further review if they included an analysis of structural imaging data involving SA or a ‘suicide-risk’ group that included suicide attempters. Reference lists of all remaining publications were examined to identify additional papers that had not been identified using the above search terms in PubMed. Any cortical or subcortical region that showed a replicated significant association with lifetime SA (demonstrated by significant effects in 2 or more publications), was selected for subsequent statistical analysis (Table S1).

**Table 1.**
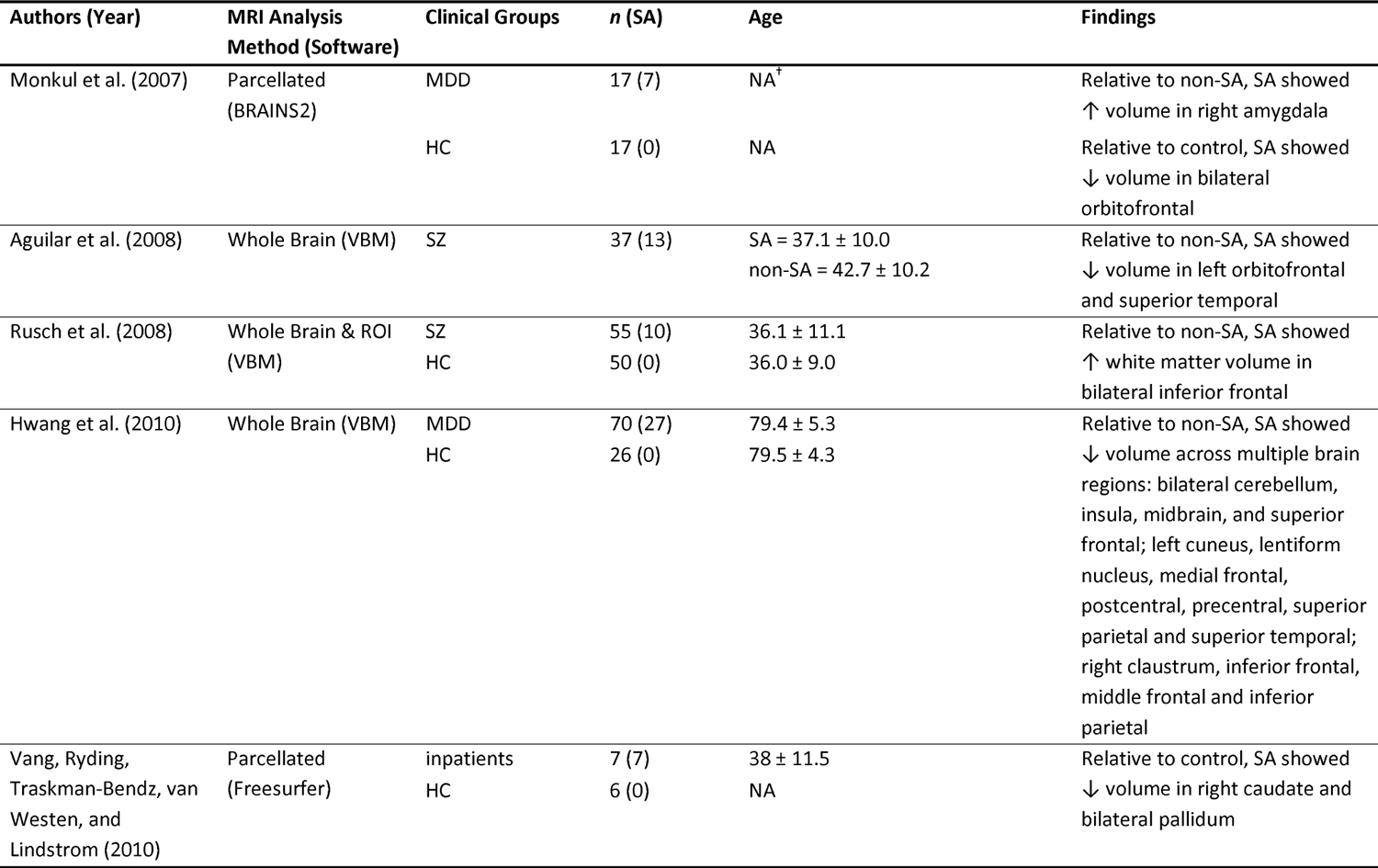
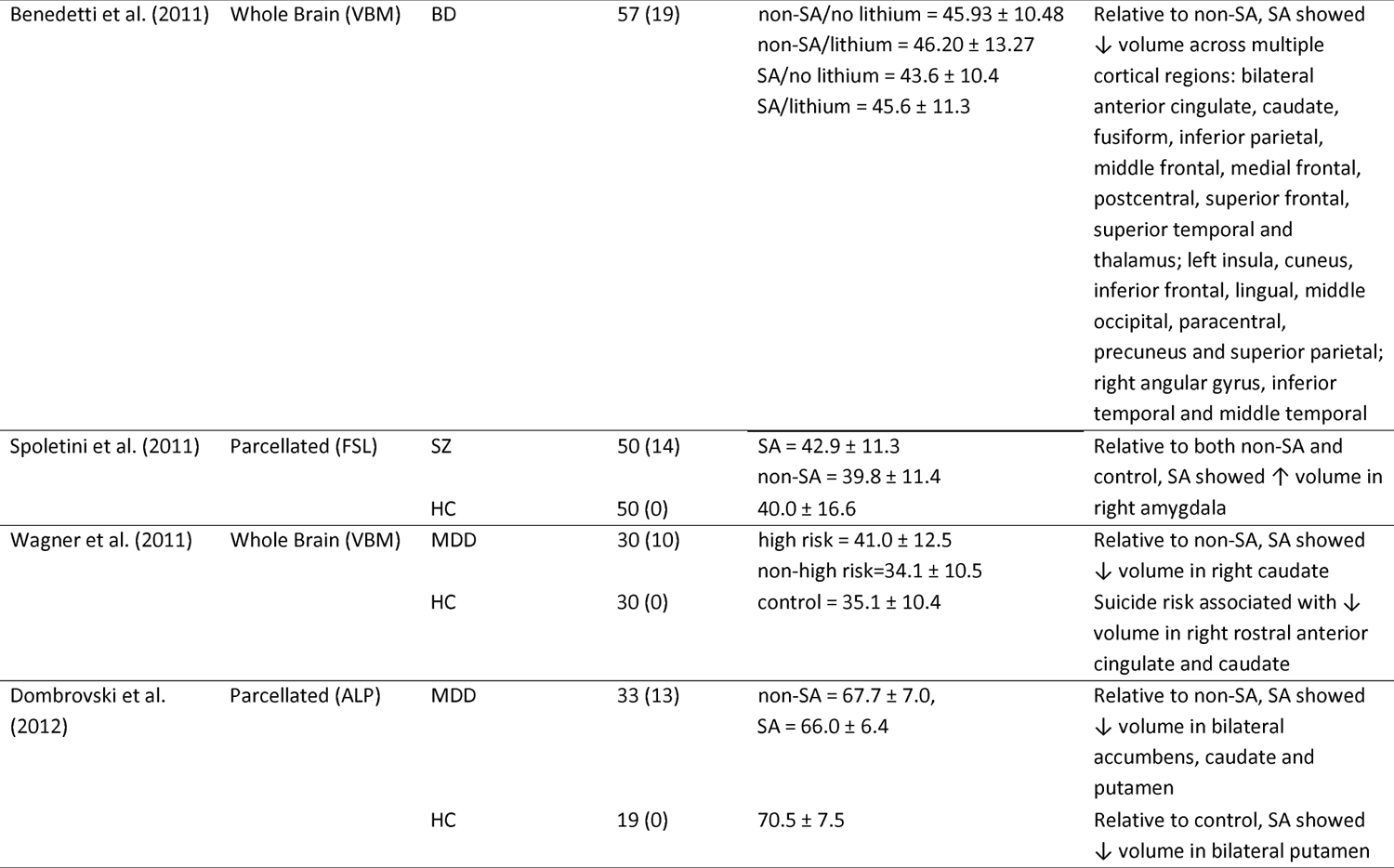
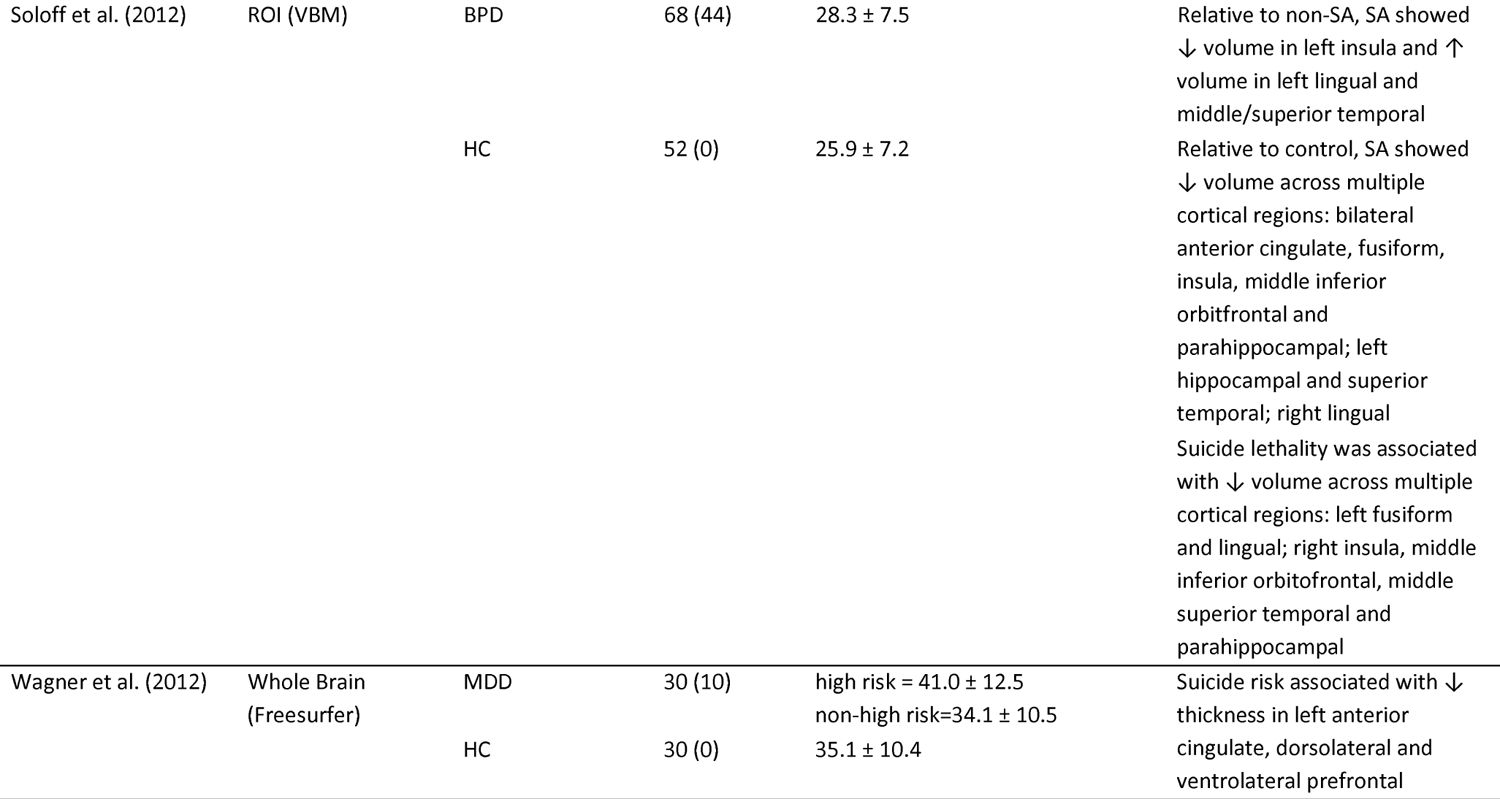
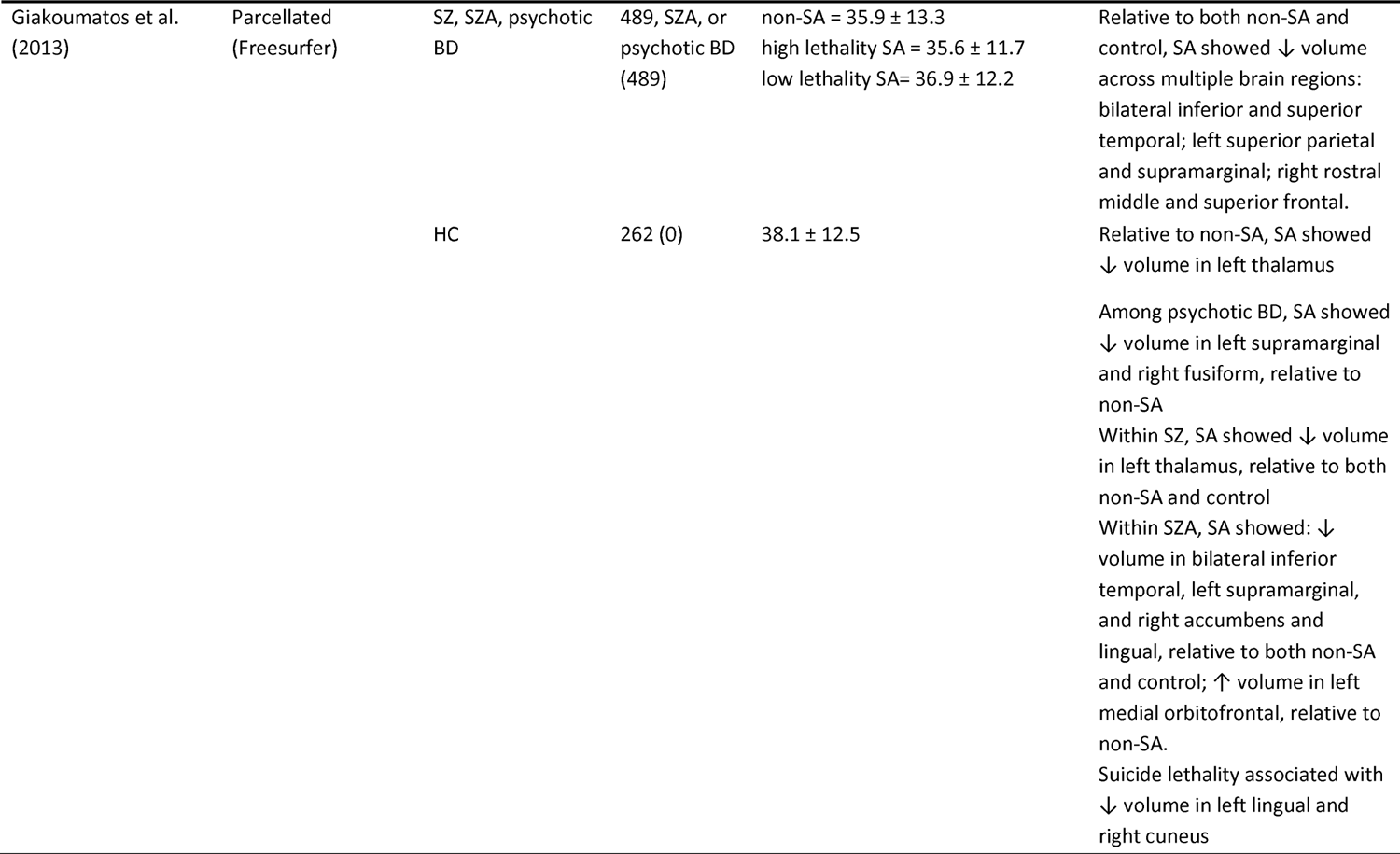
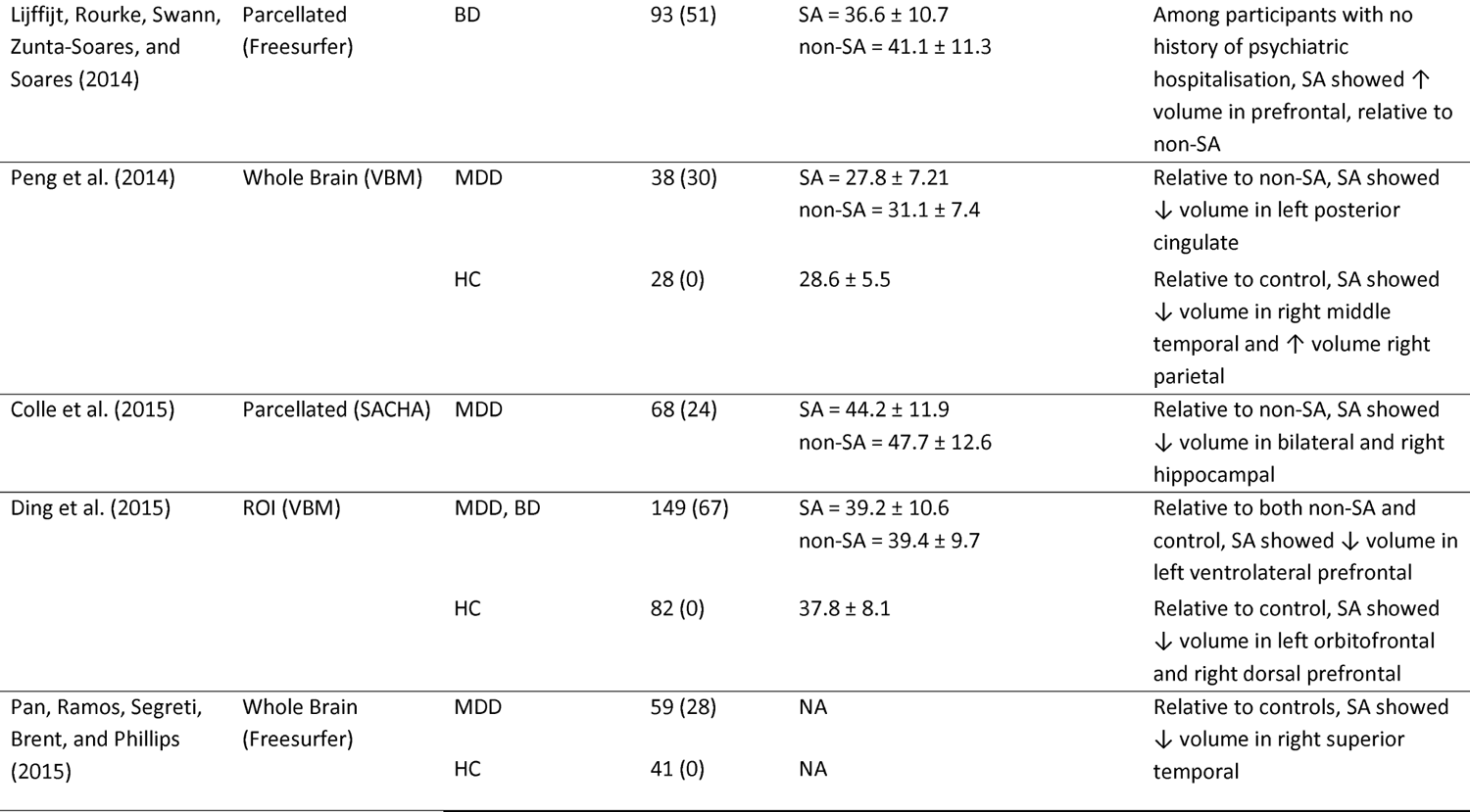
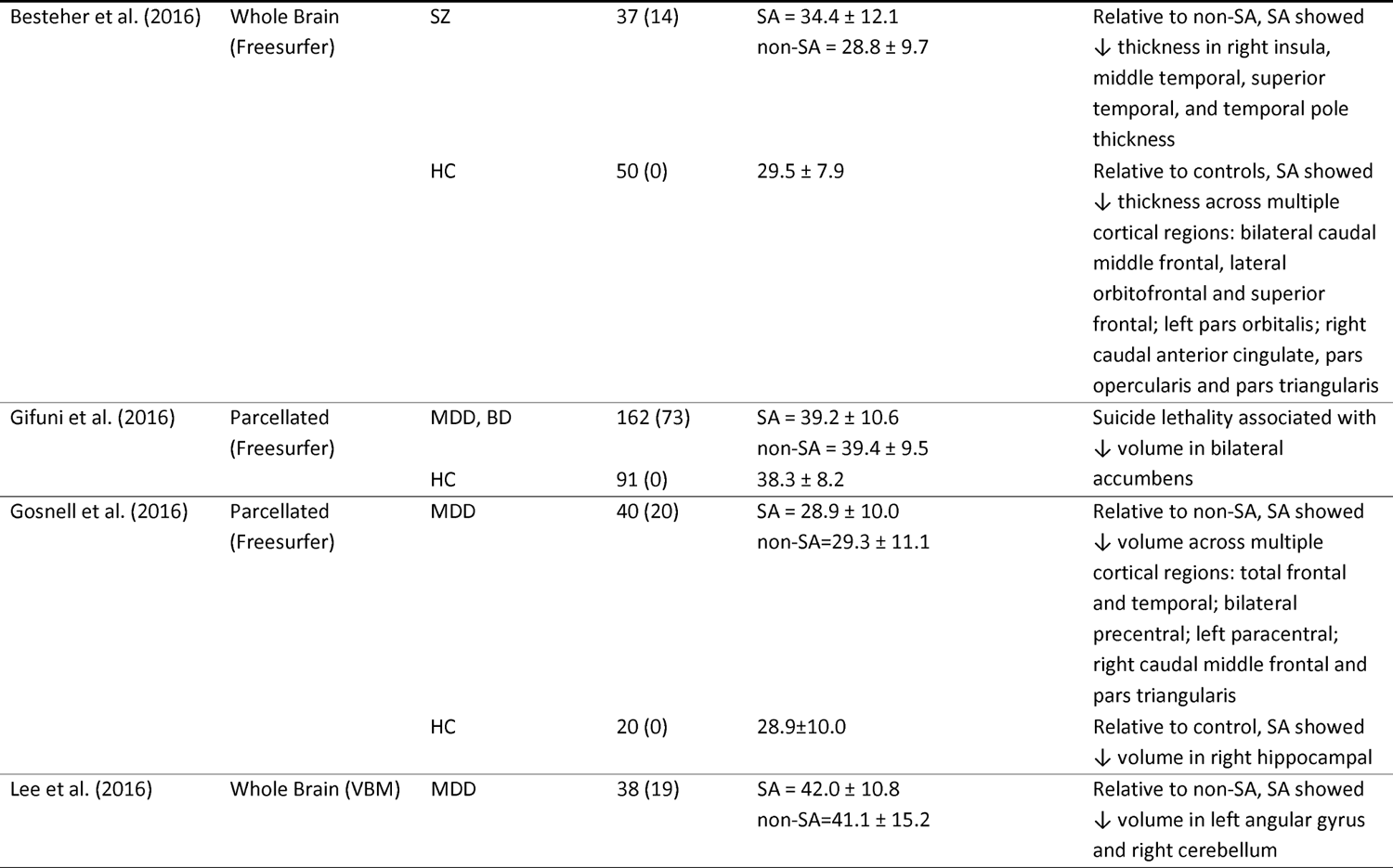
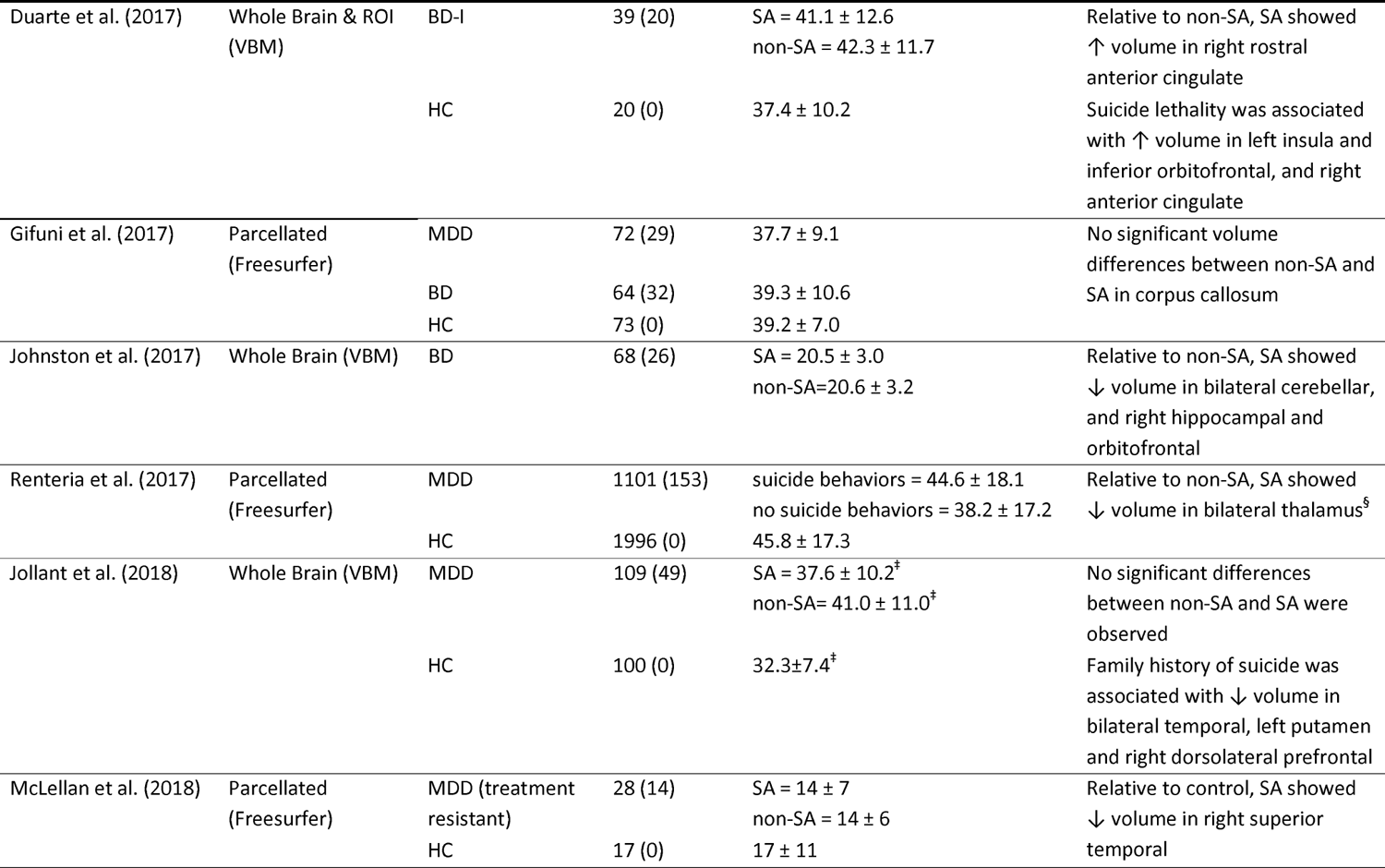
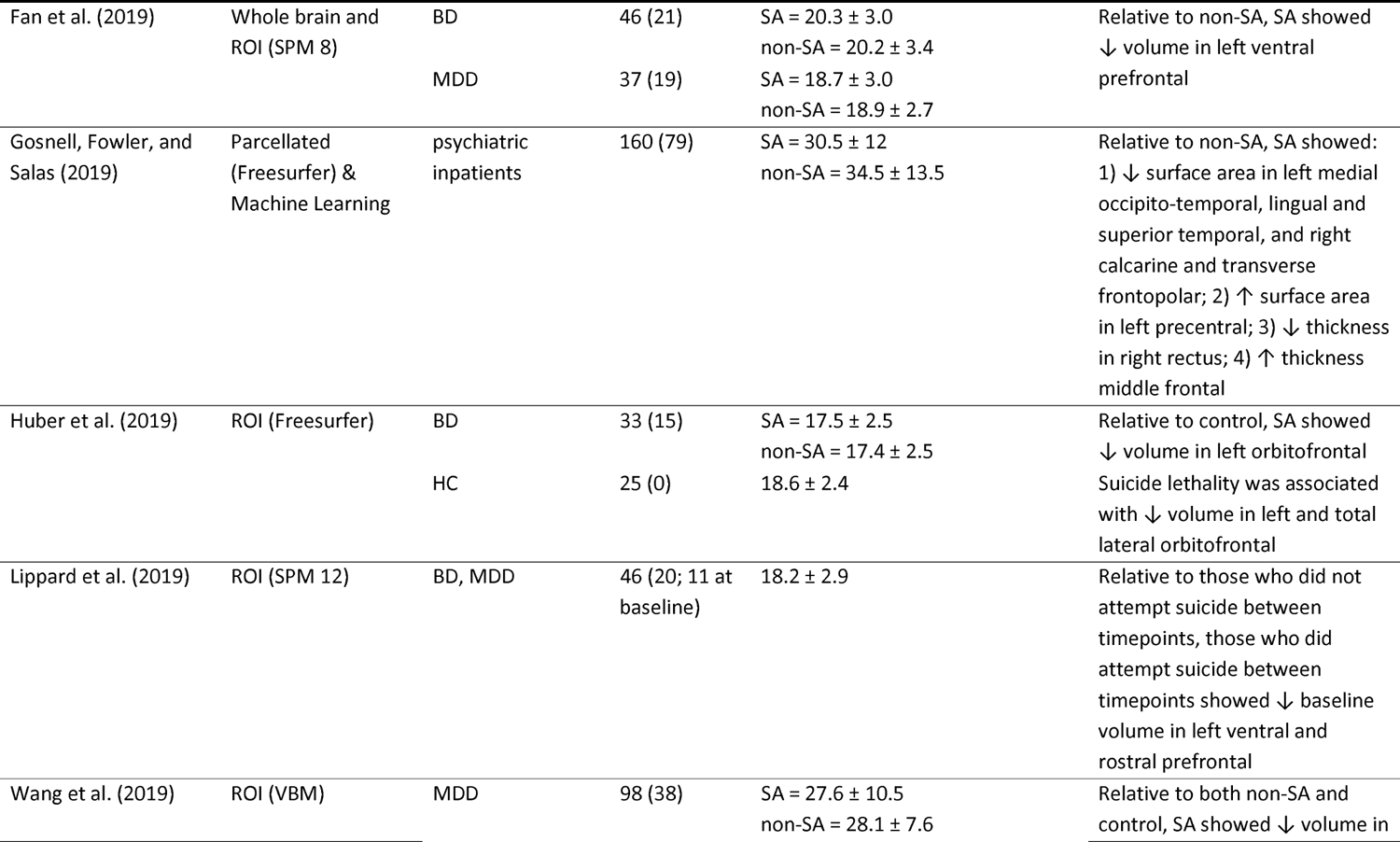
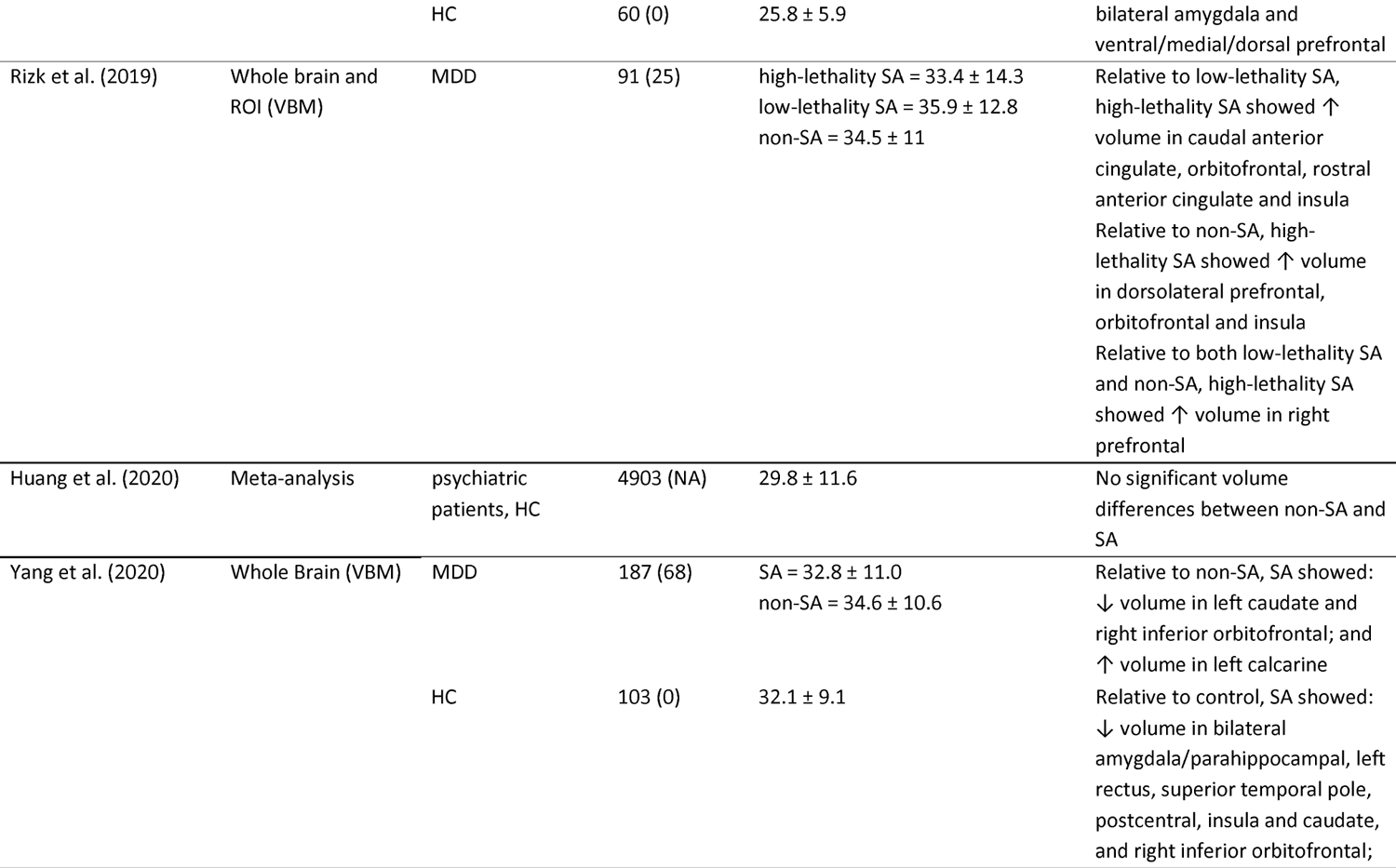
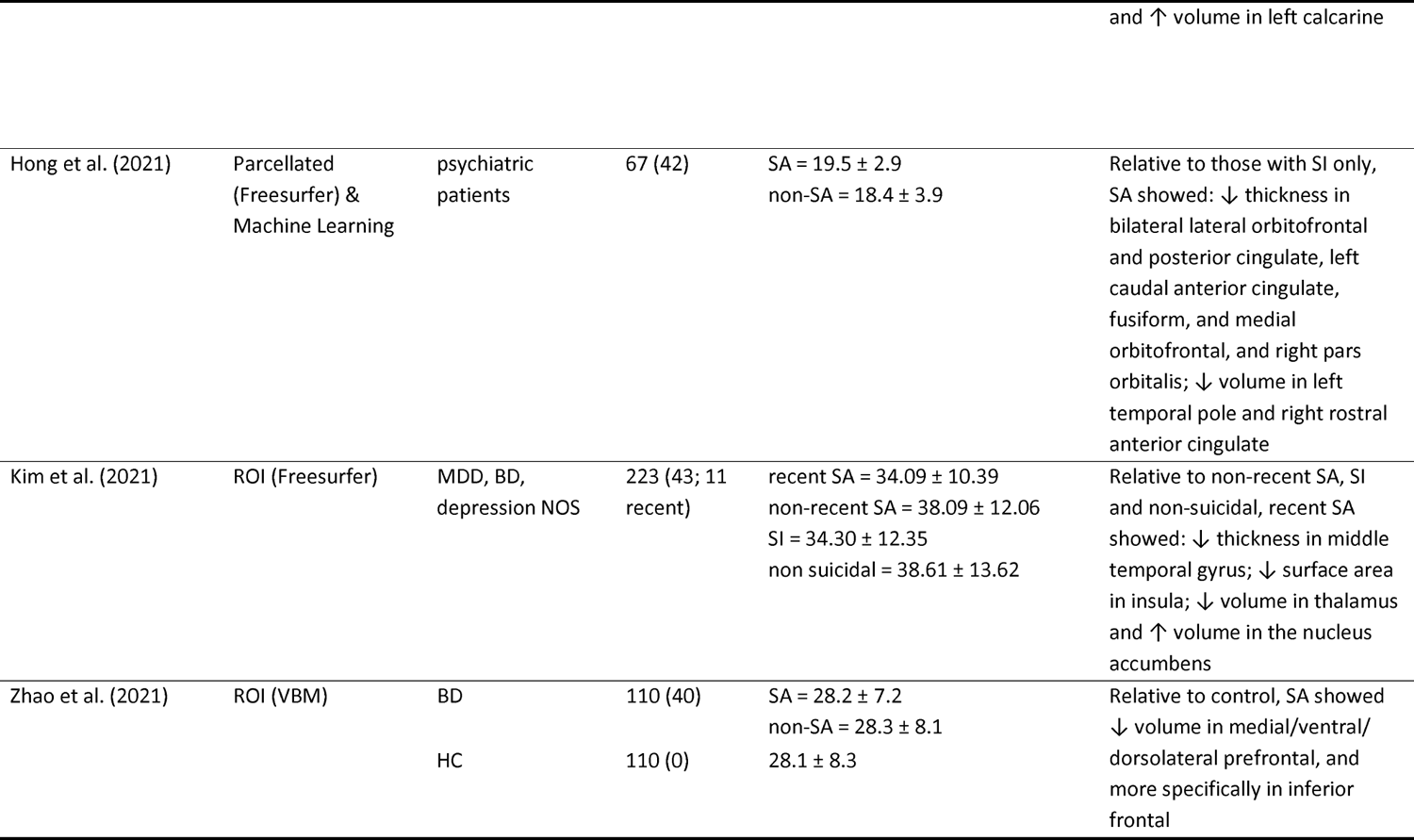
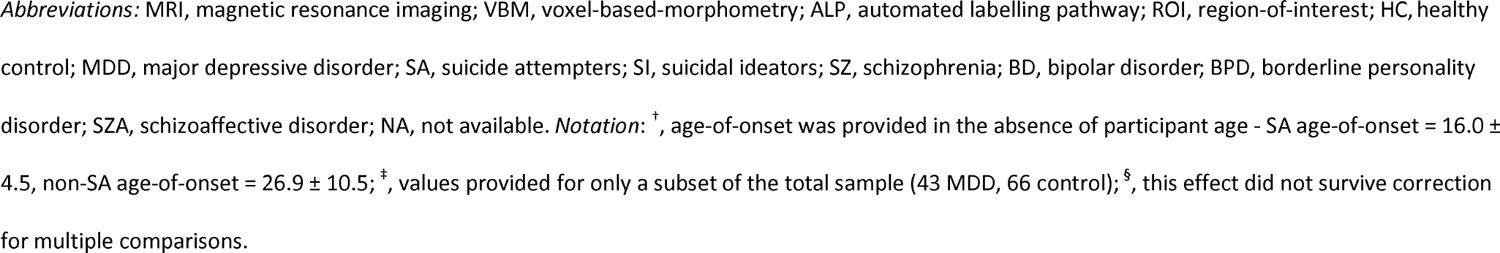
Summary of papers identified in literature review of structural imaging differences associated with suicide attempts.

## Statistical Analysis

All analyses were conducted in IBM SPSS Statistics for Windows v25 (IBM corp., Armonk, NY). None of the continuous clinical or demographic variables satisfied normality assumptions, so non-parametric Kruskal-Wallis H-tests were used to explore overall group differences. While group differences in categorical variables were assessed using Chi-square tests of independence, Fisher’s exact test of significance was used for group comparisons of lithium, antipsychotic and ‘other’ medication use, as Chi-Square assumptions were violated for these variables. All PRS were normally distributed and were analysed for group differences using ANCOVA, including the first two genetic principal components as covariates. In instances of significant group differences, post-hoc bivariate group comparisons were examined using Mann-Whitney U-tests (for non-normally distributed continuous variables), Chi-square tests (for categorical variables), and ANCOVAs (for normally distributed PRS, including the first two genetic principal components as covariates). Logistic regression analyses (across and within groups) were used to explore associations between suicidal behaviors and each PRS (including the first two genetic principal components as covariates). Relationships between PRS were examined using bivariate correlation analysis.

For each of the three PRS, the primary analysis involved construction of two independent regression models to examine associations between SA-relevant brain ROIs (as the dependent variable) and standardized PRS: 1) model 1 included standardized PRS as an independent predictor and covariates for gender and age, irrespective of group differences; 2) model 2 included standardized PRS as an independent predictor and covariates of gender, age, and group, thereby accounting for potential group differences in cortical variability. To examine the interaction effects of PRS and group on SA-relevant brain ROIs, a two-step interaction approach was utilized. Firstly, a threshold of p≤.2 was used to identify PRS main effects that demonstrated provisional evidence of a PRS-ROI association, in model 1, model 2, or both models 1 and 2. Provisional PRS-ROI associations were then examined using an additional regression model; regression model 3 included standardized PRS as an independent predictor and covariates of gender, age, and group, with the addition of model terms for the interaction between standardized PRS and group (PRS×Group), thereby accounting for potential group differences in the relationship between PRS and cortical variability. For the purposes of regression models 2 and 3, the three-level categorical group variable (BD, ‘high-risk’, control) was recoded into two dummy coded variables representing the comparisons of control vs. ‘high-risk’ (control = 0, ‘high-risk’ = 1 and BD = 0) and control vs. BD (control = 0, ‘high-risk’ = 0 and BD = 1). Therefore, the model 3 interaction terms represented the interaction of standardized PRS with these two bivariate group predictors. The dependent variable in each model represented one of the cortical or subcortical ROIs (Supplementary Table S1). Formal statistical analysis of the PRSxGroup effects in the subset of participants who had experienced SA was not performed due to insufficient sample size in control and ‘high-risk’ groups (control n = 2 and ‘high-risk’ n = 3), but participants who reported SI or SA were indicated in scatterplots for significant PRSxGroup effects. Directions of effect of PRS×Group interactions were interpreted from the relevant regression beta values, and were confirmed by visual inspection of generated scatterplots. Finally, correction for multiple comparisons was completed using false discovery rate (FDR) with a significance threshold of q<.05, accounting for the total number of tests run across all PRS, all ROIs and all regression models.

While cortical volume represents the product of surface area and thickness, exclusive examination of this three-dimensional measure may prevent the identification of unique genetic determinants of cortical structure that have been shown to be largely independent for the multiplicands of cortical area and thickness (Panizzon et al., 2009; Winkler et al., 2010; Winkler et al., 2009); therefore, all three measures (area, thickness and volume) were considered as dependent variables in independent regression models of cortical structure, whereas only volume was employed for subcortical models. Furthermore, as the model outcome (brain ROI) will be impacted by genetic admixture to a lesser extent than the PRS (particularly in this sample of European ancestry participants who were selected to be ancestrally homogeneous using MDS analysis; Supplementary Figure S1), we did not covary for genetic principal components in the primary regression models of cortical structure. However, to ensure ROI-PRS associations were not influenced by population stratification, post-hoc regression models were computed to examine all PRS on ROI effects, following the addition of principal components 1 and 2 as predictors, across all primary regression models. Post-hoc testing was also completed to examine the effects of medication use on FDR corrected significant associations between PRS and ROI. Specifically, any model that demonstrated an FDR corrected significant PRS main or interaction effect for any ROI was rerun following the addition of a medication use covariate; two binary medication use variables were computed to index either 1) use of any psychotropic medication, or 2) use of any antidepressant medication.

## Results

### Literature review of brain regions of interest (ROI) previously associated with suicide attempt

Building on the work of Cox Lippard, Johnston and Blumberg (2014), we conducted a literature review that resulted in the identification of 36 papers that reported structural imaging analysis of SA, and 2 papers that reported structural imaging analysis of a unique ‘suicide-risk’ phenotype - indexing personal history of SA and/or family history of either SA or suicide-mortality (Wagner et al., 2011; Wagner et al., 2012). A full summary of the literature review findings is detailed in Table 1; this includes significant findings relating to suicide lethality that were included as additional analyses to SA in 7 reviewed publications (Duarte et al., 2017; Giakoumatos et al., 2013; Gifuni et al., 2016; Huang, Rootes-Murdy, Bastidas, Nee, & Franklin, 2020; Huber et al., 2019; Rizk et al., 2019; Soloff et al., 2012). Subsequently, any cortical or subcortical region that showed a replicated significant association with SA (demonstrated by significant effects in 2 or more publications; Supplementary Table S1) was selected for analysis; this resulted in the selection of 24 Desikan-Killiany cortical ROIs and 8 subcortical ROIs per hemisphere (Figure 1; Supplementary Figure S2).

**Figure 1.**
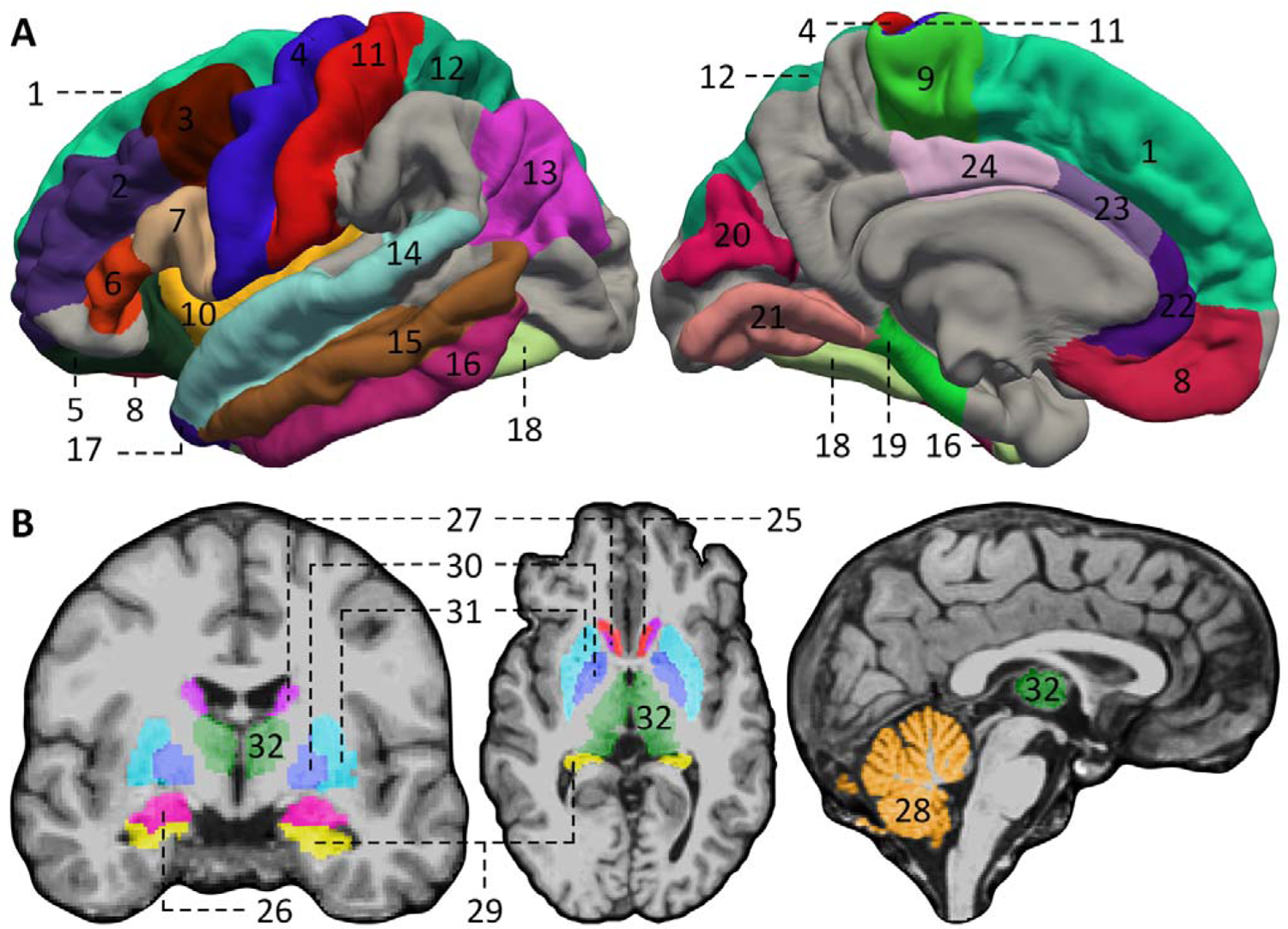
Location and boundaries of target regions-of-interest (ROIs) that were selected for analysis in this study on the basis of the literature review of structural neuroimaging studies in suicide attempt. A) cortical ROIs; 1 = superior frontal, 2 = rostral middle frontal, 3 = caudal middle frontal, 4 = precentral, 5 = lateral orbitofrontal, 6 = pars triangularis, 7 = pars opercularis, 8 = medial orbitofrontal, 9 = paracentral, 10 = insula, 11 = postcentral, 12 = superior parietal, 13 = inferior parietal, 14 = superior temporal, 15 = middle temporal, 16 = inferior temporal, 17 = temporal pole, 18 = fusiform, 19 = parahippocampal, 20 = cuneus, 21 = lingual, 22 = rostral anterior cingulate, 23 = caudal anterior cingulate, 24 = posterior cingulate. B) Subcortical ROIs: left, axial view; middle, coronal view; right, sagittal view. 25= accumbens, 26 = amygdala, 27 = caudate, 28 = cerebellum cortex, 29 = hippocampus, 30 = putamen, 31 = pallidum, 32 = thalamus proper. All brain slice images were centered at voxel 127, 127, 128 (RAS 6.4, 18.0, 1.0). A rotating image of the subcortical ROIs in three-dimensional space is provided in Supplementary Figure S2.

### Descriptive Comparisons

Groupwise comparisons of clinical, demographic and genetic variables are presented in Table 2, and post-hoc comparisons of pairwise group differences are presented in Table S2. While gender did not differ significantly between groups, the control group was on average 1.98 years older than ‘high-risk’ (uncorrected p=.023), while the BD group was 3.33 years older than control (uncorrected p<.001) and 5.32 years older than ‘high-risk’ (uncorrected p<.001). Significant group differences were observed for rates of lifetime SI (uncorrected p<.001) and SA (uncorrected p<.001), with higher reports of SI and SA in the BD group when compared to both control and ‘high-risk’ groups, and higher SI in ‘high-risk’ when compared to controls. Significant group differences were also observed for lifetime DSM-IV diagnosis, with higher rates of at least one major depressive episode (MDE), recurrent MDD, anxiety and substance disorders in BD, relative to both controls and ‘high-risk’, and higher rates of at least one MDE, recurrent MDD and anxiety disorders in ‘high-risk’, relative to controls. No group differences were observed for current MDE and hypo/manic episodes. Comparison of symptom severity scales revealed higher MADRS and CDI in BD, relative to both controls and ‘high-risk’, and higher MADRS in ‘high-risk’, relative to controls. While higher rates of all types of psychiatric medication use were observed in BD, relative to both controls and ‘high-risk’, no group differences were observed in medication use between controls and ‘high-risk’, and the use of mood stabilisers, lithium and antipsychotics was not observed for any control or ‘high-risk’ participant. Significant group differences were also observed in the PRS for SA-in-BD (F=4.482, uncorrected p=.012; Table 2), with control participants showing significantly lower mean PRS when compared with ‘high-risk’ (-.245 versus .114, uncorrected p=.006) and BD (-.245 versus .145, uncorrected p=.009) (Table S2). While ‘high-risk’ demonstrated a slightly lower mean PRS for SA-in-BD when compared to BD (.114 versus .145, respectively; Table 2), this difference was not significant (uncorrected p=.852). No group differences were observed for SA-in-MDD PRS or Risky Behavior PRS. Bivariate correlation analysis of each PRS indicated a negligible non-significant relationship between SA-in-BD and SA-in-MDD PRS (r=.092), and negligible non-significant relationship of each SA PRS with Risky Behavior (r=.069 and .087, respectively). Finally, no significant associations were observed between suicide behaviors (lifetime SI or SA) and any PRS.

**Table 2.**
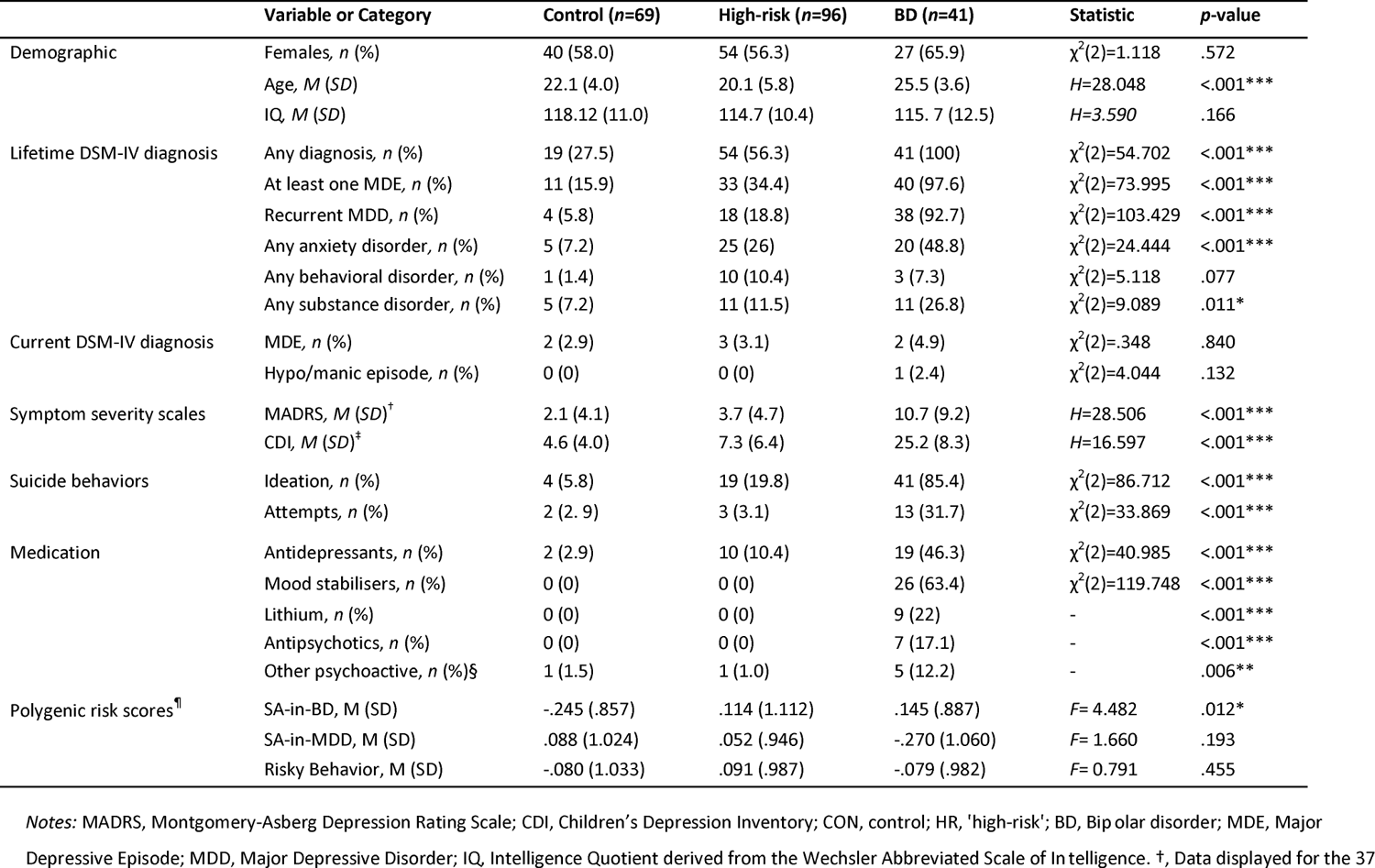
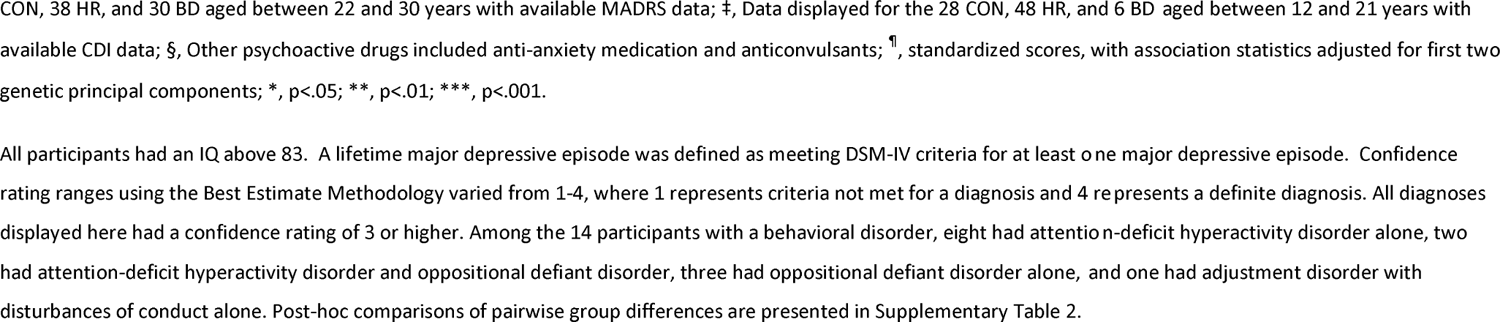
Summary statistics and group comparisons for demographic, clinical, and PRS^2^ variables.

|  | Variable or Category | Control (n=69) | High-risk (n=96) | BD (n=41) | Statistic | p-value |
| --- | --- | --- | --- | --- | --- | --- |
| Demographic | Females, n (%) | 40 (58.0) | 54 (56.3) | 27 (65.9) | $\chi^2(2)=1.118$ | .572 |
| | Age, M (SD) | 22.1 (4.0) | 20.1 (5.8) | 25.5 (3.6) | $H=28.048$ | <.001*** |
| | IQ, M (SD) | 118.12 (11.0) | 114.7 (10.4) | 115.7 (12.5) | $H=3.590$ | .166 |
| Lifetime DSM-IV diagnosis | Any diagnosis, n (%) | 19 (27.5) | 54 (56.3) | 41 (100) | $\chi^2(2)=54.702$ | <.001*** |
| | At least one MDE, n (%) | 11 (15.9) | 33 (34.4) | 40 (97.6) | $\chi^2(2)=73.995$ | <.001*** |
| | Recurrent MDD, n (%) | 4 (5.8) | 18 (18.8) | 38 (92.7) | $\chi^2(2)=103.429$ | <.001*** |
| | Any anxiety disorder, n (%) | 5 (7.2) | 25 (26) | 20 (48.8) | $\chi^2(2)=24.444$ | <.001*** |
| | Any behavioral disorder, n (%) | 1 (1.4) | 10 (10.4) | 3 (7.3) | $\chi^2(2)=5.118$ | .077 |
| | Any substance disorder, n (%) | 5 (7.2) | 11 (11.5) | 11 (26.8) | $\chi^2(2)=9.089$ | .011* |
| Current DSM-IV diagnosis | MDE, n (%) | 2 (2.9) | 3 (3.1) | 2 (4.9) | $\chi^2(2)=.348$ | .840 |
| | Hypo/manic episode, n (%) | 0 (0) | 0 (0) | 1 (2.4) | $\chi^2(2)=4.044$ | .132 |
| Symptom severity scales | MADRS, M (SD)<sup>†</sup> | 2.1 (4.1) | 3.7 (4.7) | 10.7 (9.2) | $H=28.506$ | <.001*** |
| | CDI, M (SD)<sup>†</sup> | 4.6 (4.0) | 7.3 (6.4) | 25.2 (8.3) | $H=16.597$ | <.001*** |
| Suicide behaviors | Ideation, n (%) | 4 (5.8) | 19 (19.8) | 41 (85.4) | $\chi^2(2)=86.712$ | <.001*** |
| | Attempts, n (%) | 2 (2.9) | 3 (3.1) | 13 (31.7) | $\chi^2(2)=33.869$ | <.001*** |
| Medication | Antidepressants, n (%) | 2 (2.9) | 10 (10.4) | 19 (46.3) | $\chi^2(2)=40.985$ | <.001*** |
| | Mood stabilisers, n (%) | 0 (0) | 0 (0) | 26 (63.4) | $\chi^2(2)=119.748$ | <.001*** |
|  | Lithium, n (%) | 0 (0) | 0 (0) | 9 (22) | - | <.001*** |
|  | Antipsychotics, n (%) | 0 (0) | 0 (0) | 7 (17.1) | - | <.001*** |
|  | Other psychoactive, n (%)§ | 1 (1.5) | 1 (1.0) | 5 (12.2) | - | .006** |
| Polygenic risk scores<sup>¶</sup> | SA-in-BD, M (SD) | -.245 (.857) | .114 (1.112) | .145 (.887) | $F=4.482$ | .012* |
| | SA-in-MDD, M (SD) | .088 (1.024) | .052 (.946) | -.270 (1.060) | $F=1.660$ | .193 |
| | Risky Behavior, M (SD) | -.080 (1.033) | .091 (.987) | -.079 (.982) | $F=0.791$ | .455 |

Notes: MADRS, Montgomery-Asberg Depression Rating Scale; CDI, Children's Depression Inventory; CON, control; HR, 'high-risk'; BD, Bipolar disorder; MDE, Major Depressive Episode; MDD, Major Depressive Disorder; IQ, Intelligence Quotient derived from the Wechsler Abbreviated Scale of Intelligence. †, Data displayed for the 37
CON, 38 HR, and 30 BD aged between 22 and 30 years with available MADRS data; ‡, Data displayed for the 28 CON, 48 HR, and 6 BD aged between 12 and 21 years with available CDI data; §, Other psychoactive drugs included anti-anxiety medication and anticonvulsants; ¶, standardized scores, with association statistics adjusted for first two genetic principal components; \*, $p < .05$ ; \*\*, $p < .01$ ; \*\*\*, $p < .001$ .

### PRS and ROI associations

A full list of uncorrected and FDR corrected ROI findings for primary regression models 1, 2, and 3, are presented in supplementary Table S2. Of the 480 PRS and ROI associations that were examined in regression models 1 and 2, 81 associations were selected using the two-step interaction approach, for examination of PRS and group interaction effects using regression model 3.

### Effects of PRS for SA in BD

After FDR correction, models 1 and 2 revealed a significant main effect of SA-in-BD PRS on right rostral anterior cingulate volume (Table 3), controlling for age and gender, and both before and after controlling for group membership. Specifically, higher PRS was associated with greater right rostral anterior cingulate volume. After also controlling for potential PRS×Group interactions in model 3, this PRS main effect was not significant. However, model 3 did reveal FDR corrected significant main effects of PRS on left parahippocampal thickness and volume, controlling for age, gender, group, and potential PRS×Group interactions (Table 4). Alongside significant main effects of PRS, significant PRS×Group interactions were also observed in model 3 for left parahippocampal thickness and volume. However, only the PRS×Group (Control vs BD) interaction on left parahippocampal volume survived FDR correction. While greater left parahippocampal volume was observed in controls with lower PRS, the inverse relationship was seen in BD, with greater volume associated with higher PRS (Figure 2A); on average and relative to controls, for each standard deviation unit increase in the SA-in-BD PRS, BD participants demonstrated additional left parahippocampal volume of .220cm^3^. All FDR corrected significant SA-in-BD PRS main and interaction effects from models 1 to 3 were also observed in post-hoc analyses that controlled for genetic principal components 1 and 2 (Table S3) and ‘any psychotropic’ or ‘any antidepressant’ medication use (Table S4). Visual inspection of PRS×Group interaction plots displaying the subpopulations of suicidal ideators and attempters did not reveal any patterns of association between either type of suicide-related behavior and any ROI or the SA-in-BD PRS (Figure 2A).

**Figure 2.**
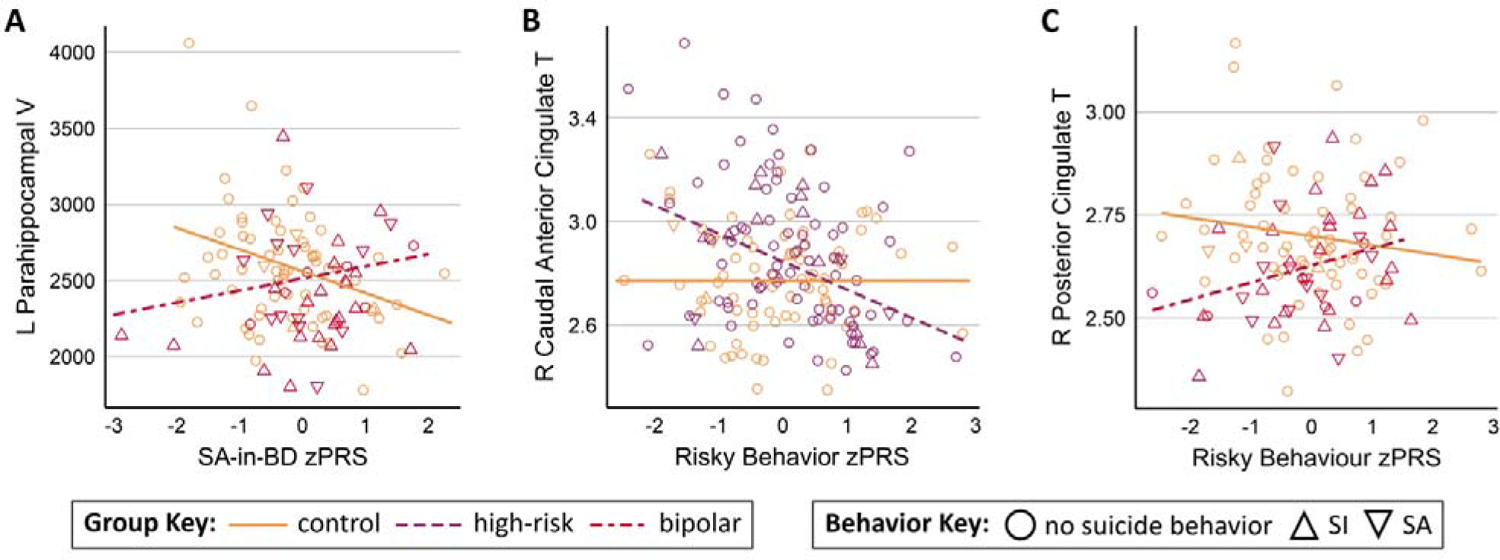
Scatterplots displaying significant FDR corrected PRS×Group interaction effects for linear regression model 3 (covarying for age, gender and group). Participants who reported suicide-behaviors during clinical interview are indicated with triangle data points (SI up arrow and SA down arrow) and those with no suicide behavior are indicated with circles. Panels A and C show FDR corrected control (orange) vs. BD (magenta) comparisons. Panel B shows FDR corrected control (orange) vs. ‘high-risk’ (purple) comparisons. Abbreviations: FDR, survived false discovery rate; SI, suicide ideation; SA, suicide attempts; zPRS, standardized polygenic risk score; BD, bipolar disorder; V, Volume; T, Thickness.

**Table 3.**
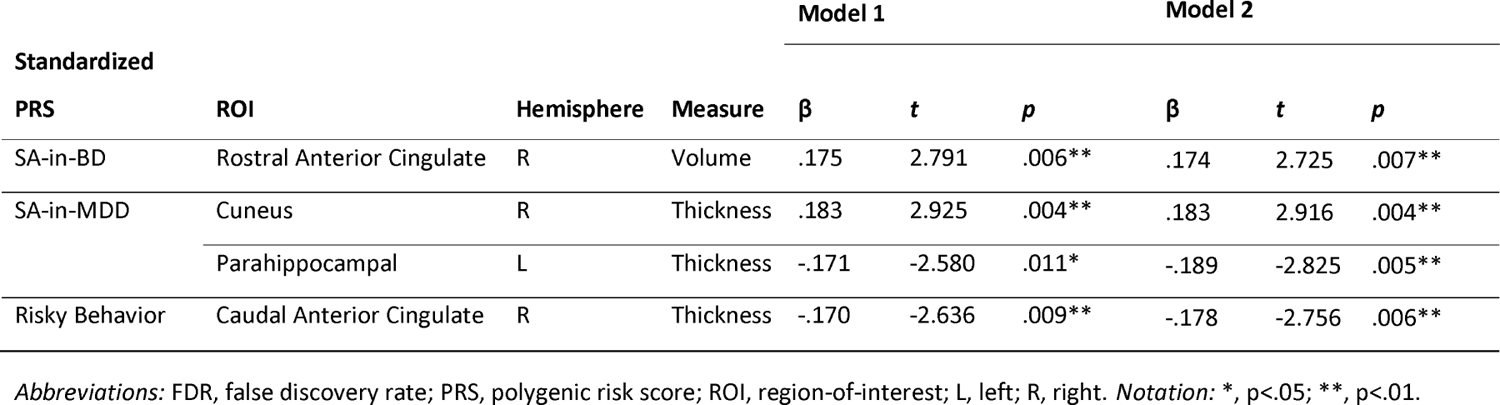
FDR corrected significant main effects for each independent polygenic risk score in models 1 (irrespective of group) & 2 (covarying for group), covarying for age and gender.

### Effects of PRS for SA in MDD

After FDR correction, models 1 and 2 revealed significant main effects of SA-in-MDD PRS on left parahippocampal and right cuneus thickness (Table 3), controlling for age and gender, and both before and after controlling for group membership. While higher PRS was associated with thicker cortex in the right cuneus, higher PRS associated with thinner cortex in the left parahippocampal region. These effects remained significant in post-hoc models 1 and 2 that controlled for genetic principal components 1 and 2 (Table S3) and ‘any psychotropic’ or ‘any antidepressant’ medication use (Table S4). When examining potential PRS×Group interactions in model 3, these PRS main effects and corresponding PRS×Group interaction terms were not significant after FDR correction.

### Effects of PRS for Risky Behavior

After FDR correction, models 1 and 2 revealed a significant main effect of the Risky Behavior PRS on right caudal anterior cingulate thickness (Table 3), controlling for age and gender, and both before and after controlling for group membership. Specifically, higher PRS was associated with thinner cortex. After also controlling for potential PRS×Group interactions in model 3, this PRS main effect was not significant. However, the PRS×Group (Control vs ‘high-risk’) interaction term was significant in this model (after FDR correction), with thicker right caudal anterior cingulate cortex in ‘high-risk’ with lower PRS, and almost no relationship between PRS and cortical thickness in controls (Figure 2B); on average and relative to control, for each standard deviation unit increase in the Risky Behavior PRS, ‘high-risk’ participants demonstrated an additional .299cm lower right caudal anterior cingulate thickness. After FDR correction, model 3 also revealed a significant PRS×Group (Control vs BD) interaction term for right posterior cingulate thickness. While thicker right posterior cingulate cortex was observed in controls with lower PRS, the inverse relationship was seen in BD, with thicker cortex associated with higher PRS (Figure 2C); on average and relative to controls, for each standard deviation unit increase in the Risky Behavior PRS, BD participants demonstrated additional right posterior cingulate thickness of .178cm. All FDR corrected significant Risky Behavior PRS main and interaction effects from models 1 to 3 were also observed in post-hoc analyses that controlled for genetic principal components 1 and 2 (Table S3) and ‘any psychotropic’ or ‘any antidepressant’ medication use (Table S4). Visual inspection of PRS×Group interaction plots displaying the subpopulations of suicidal ideators and attempters did not reveal any patterns of association between either type of suicide-related behavior and any ROI or the risky behavior PRS (Figure 2B-C).

## Discussion

Despite advances in diagnosing and treating mental health disorders, suicide continues to be a major source of mortality associated with psychopathology, and is the second leading cause of death for 10-34 year olds in the USA (Centers for Disease Control and Prevention, February 19, 2017) and the leading cause of death in 15-44 year olds in Australia (Australian Institute of Health and Welfare, 2021). Among youth with mental health symptoms, some never attempt suicide, yet for others, suicidal behavior is pervasive and recurrent; thus predictive models that incorporate a comprehensive set of variables, including biological data, are sorely needed (Franklin et al., 2017). Recent gene discovery efforts to define genomic signatures of SA within the context of psychopathology (Mullins et al., 2019) have presented opportunities to explore relationships between suicide-specific genomic profiles and cortical variability, as viable biomarkers with potential for clinical utility. The present study detailed an exploratory analysis of the effects of PRS for SA and risky behavior on structural variance in cortical ROIs that have previously shown replicable associations with SA.

In the present study, the anterior cingulate demonstrated significant associations with two independent PRS. Firstly, a significant positive association was observed between the SA-in-BD PRS and right rostral anterior cingulate volume, both before and after controlling for clinical group membership. Secondly, a significant positive association between the Risky Behavior PRS and right caudal anterior cingulate thickness was observed to be modified by familial risk for BD; thinner right caudal anterior cingulate was observed at higher PRS in ‘high-risk’, while the control group showed no PRS and caudal anterior cingulate association. The anterior cingulate has repeatedly been identified in previous suicide imaging literature, with smaller volume associated with SA in patients with BD (Benedetti et al., 2011) and borderline personality disorder (Soloff et al., 2012), suicide risk in MDD (Wagner et al., 2011; Wagner et al., 2012), and suicide lethality in schizophrenia, schizoaffective disorder, and BD (Giakoumatos et al., 2013). A thinner anterior cingulate cortex has also been associated with SA in schizophrenia (Besteher et al., 2016) and suicide risk in MDD (Wagner et al., 2011; Wagner et al., 2012). In contrast, a single study of patients with BD type-I observed an association between larger anterior cingulate volume and both SA and lethality (Duarte et al., 2017); a finding that has not been replicated to date. Functionally, the rostral section of the anterior cingulate has been associated with limbic, autonomic, memory and reward processes, with projections into the amygdala, lateral hypothalamus, hippocampus, orbitofrontal and ventral striatum (Stevens, Hurley, & Taber, 2011). Functional imaging studies have highlighted the role of this region in emotional learning, assessment and regulation, which is of particular relevance to the emotional dysregulation observed in BD (Townsend & Altshuler, 2012) and the well-established links between emotional dysregulation and suicidal behavior (Neacsiu, Fang, Rodriguez, & Rosenthal, 2018; Rajappa, Gallagher, & Miranda, 2012). In contrast, the caudal portion of the anterior cingulate has been associated with cognitive functions including executive control and conflict monitoring, with extensive connections to the lateral prefrontal and motor cortices (Stevens et al., 2011). In support of the present association between the Risky Behavior PRS and caudal anterior cingulate structure in ‘high-risk’, previous research has established that deficits in executive function account for a substantial proportion of the variance in risky behavior (Reynolds, Basso, Miller, Whiteside, & Combs, 2019) and that deficits in executive function are observed in first-degree relatives of BD probands (Arts, Jabben, Krabbendam, & van Os, 2008; Nicol Ferrier, Chowdhury, Thompson, Watson, & Young, 2004). Given the divergent functional association of the rostral and caudal anterior cingulate, as well as the present findings of significant associations between these ROIs and two uncorrelated PRS that index separate behavioral phenotypes (SA and risky behavior), it is possible that structural changes in these anterior cingulate subdivisions relate to differential processes that contribute to suicide attempts; specifically, emotional dysregulation and engagement in risky behavior may respectively be associated with rostral anterior cingulate and caudal anterior cingulate structure.

Furthermore, the two PRS and anterior cingulate associations observed in the present studies demonstrated divergent directions of effect; greater right rostral anterior cingulate area and volume were observed at higher SA-in-BD PRS, while smaller caudal anterior cingulate thickness was observed at higher Risky Behavior PRS. While these differential associations may relate to the separate functional roles of these regions or the separate cortical measurement modalities (area vs. thickness) and their independent genetic determinants (Panizzon et al., 2009; Winkler et al., 2010; Winkler et al., 2009), it is possible that the genetic determinants of suicide risk and phenotypes that contributed to suicide risk (e.g. risky behavior) are at least partially independent, resulting in unique associations with cortical structure. Partial support for this interpretation is provided by the negligible correlation that was observed in the present study between the SA-in-BD and Risky Behavior PRS. However, this lack of PRS correlation may be due, in part, to insufficient power of the discovery GWAS to accurately index the full spectrum of genomic variants that influence suicidal behaviors and related phenotypes, despite using the largest studies currently available. It may be that integration of multiple suicide-related phenotypes in a multi-trait GWAS analysis (e.g. Turley et al., 2018), in addition to increased sample size of primary GWAS, will improve genetic prediction models.

The consistent cross-disorder finding of PRS effects on parahippocampal structure in the present study (SA-in-MDD for models 1 & 2, and SA-in-BD for model 3) indicates convergence of genetic effects for SA in this region. Structural differences in this region have been observed in neuroimaging studies of SA in MDD (Yang et al., 2020) and borderline personality disorder (Soloff et al., 2012), with smaller cortical volume observed in suicide attempters, concordant with the smaller cortical volume and thinner cortex observed here at higher PRS. In addition, resting state functional connectivity studies have identified patterns of increased connectivity between the left parahippocampus and habenula (Ambrosi et al., 2019), and decreased connectivity between the right parahippocampus and amygdala (Kang et al., 2017), in the presence of suicide behavior in MDD. While the parahippocampal gyrus has primarily been associated with visuospatial processing and the encoding of episodic memory (Aminoff, Kveraga, & Bar, 2013), the contribution of this region to cognitive valuation of rewards and prospection may be key to the observed association between parahippocampal structure and suicide attempts (Vanyukov et al., 2016); specifically, structural and associated functional deficits in the parahippocampal cortex may impair the valuation of future rewards and the assessment of immediate deterrents, increasing the risk of suicide behaviors. Support for this theory is provided by the findings of Vanyukov et al. (2016), who observed parahippocampal deactivation associated with the valuation of a delayed reward in MDD with a history of SA. While multimodal integration of genetic, clinical and imaging phenotypes of SA is beyond the scope of the present study, the observed association between polygenic risk for SA (in both MDD and BD) and parahippocampal structure provides justification for further exploration of parahippocampal deficits as a biomarker for SA across psychiatric disorders.

In the present study, a significant association was also observed between the SA-in-MDD PRS and right cuneus volume. Previous neuroimaging studies in SA have observed smaller left cuneus volume in association with SA in both BD and MDD (Hwang et al., 2010). However, in the present study, higher SA-in-MDD PRS was associated with larger right cuneus volume (before and after accounting for group). This apparent disparity in the observed direction of effect may be ascribed by several factors, including incomplete indexing of SA by the PRS, and potentially tissue-specific expression driven by genomic variants with the largest effect sizes, represented by the PRS signature. Furthermore, the higher rate of SA in previous inpatient samples (> 33%) (Benedetti et al., 2011; Hwang et al., 2010), compared to the relatively low rate of SA observed in the present ‘high-risk’ group (3%), which was not ascertained for suicide behaviors, may have obscured apparent effects. In addition, substantial age differences exist between the previous mature and elderly adult samples (Benedetti et al., 2011; Hwang et al., 2010), relative to the present adolescent and young adult sample; this may be important, as the examination of neuroimaging phenotypes at different developmental stages may result in apparent inconsistencies of effect direction, both across and within groups, related to cohort age – the cuneus reaches peak cortical thickness at ∼9 years of age (Shaw et al., 2008), followed by a quadratic decline in area, thickness and volume with increasing age (Vijayakumar et al., 2016), and the cingulate cortex reaches peak cortical thickness at ∼10.5 years of age (Shaw et al., 2008). Indeed, we recently found that young HR have greater variability in caudal anterior cingulate volume at baseline and accelerated volume reductions over time, compared to controls, over a 2-year inter-scan period(Roberts et al., 2020). It may be possible that differential developmental trajectories impacted the ROI-PRS associations reported herein. While our statistical models all included age as a covariate, only linear effects of age were modelled and PRS effects on developmental trajectories over time were not able to be assessed in this cross-sectional analysis framework.

Comprising a large proportion of the visual cortex, the cuneus has primarily been associated with basic visual processing including motion, orientation, direction and speed (Grill-Spector & Malach, 2004). The cuneus has also been shown to be involved in response inhibition functions (Matthews, Simmons, Arce, & Paulus, 2005), and deficits in response inhibition have been associated with larger right cuneus volume in BD (Haldane, Cunningham, Androutsos, & Frangou, 2008). Additionally, a history of SA in mood disorder with psychosis has been associated with decreased cuneus activation during performance of a goal-representation task, tapping into processes of cognitive control (Minzenberg et al., 2015). Finally, the cuneus has been identified as one of several regions with lower global brain connectivity associated with SA in BD (Cheng et al., 2020). These converging lines of evidence suggest that structural, functional and connectivity deficits in the cuneus, associated with mood disorder diagnoses, may contribute to deficits in cognitive control, which have been associated with increased risk of SA (Keilp et al., 2001; Richard-Devantoy, Ding, Lepage, Turecki, & Jollant, 2016; Richard-Devantoy et al., 2012). Further, the present association between cuneus structure and the SA-in-MDD PRS alludes to the possibility of a gene-brain-behavior relationship, where genetic markers for suicide risk influence cuneus structure, which may then precipitate deficits in cognitive control and subsequently increase the risk of suicidal behaviors, although this hypothesis requires formal testing in the future.

### Limitations and Future Directions

The PRS employed here were calculated using common DNA variants – as a genomic index of risk – but rare genetic variants and DNA methylation signatures also contribute to SA, and these other important sources of genomic variability were not represented in the PRS employed here. In addition, the PRS for the present study were selected to index a subset of suicide-related phenotypes but were not exhaustive – they did not index depression symptoms, anxiety or impulsive aggression, which may further exacerbate suicidal behaviors and impact cortical structure. While it could be argued that the PGC analysis of SA may not presently be powered to find disorder-specific variants, the inclusion of a PRS derived from the meta-analysis of the SA-in-BIP and SA-in-MDD GWAS (i.e. the SA-in-mood-disorder PRS) was not included herein due to its non-independence from the PRS of primary focus (i.e. the SA-in-BD PRS).. As seen in GWAS of other psychiatric disorders, the number of significant genetic associations and the proportion of phenotypic variance explained is expected to grow with increased sample sizes. Therefore, future studies employing PRS based on larger SA discovery GWAS, and potentially including multi-trait GWAS approaches using correlated suicide-related phenotypes or cross-disorder signatures, may provide invaluable biological insights beyond those studied here, which explain less than 0.2% of variance on the liability scale for SA. Finally, the molecular mechanisms underlying SI and SA may be distinct, so examination of biomarkers for both suicide-related phenotypes is warranted.

In relation to the brain ROI that were examined, this study focused on regions that were implicated in cross-sectional studies of lifetime suicide attempt, thus brain changes that occur proximal to attempt events may not have been captured or examined. In addition, as participants in this study were drawn from an ongoing longitudinal study of familial risk for BD, subjects were not ascertained by the presence of suicide behaviors during participant recruitment. Therefore, the number of attempters was too small to enable robust analysis of PRS effects on SA. We also note that clinical identification of SA has a strong reliance on self-report, which may lead to underestimation of suicidal behavior. In addition, it is also possible that use of specific psychotropic medications (other than antidepressants) in the present sample influenced brain structure in the ROIs that were examined. However, psychotropic medication use (other than antidepressants) was observed exclusively in the BD group and not observed in the control and ‘high-risk’ groups. Further, sample size limitations in the BD group prevented stratification of brain structure by medication type.

As this study is longitudinal in nature, it may be possible to model future SA with adequate biomarkers, and model associations between longitudinal neuroimaging trajectories and polygenic variability associated with SA. Future prospective studies that include a broad range of biological, environmental and experiential constructs and their interactions over the course of early development are needed (Turecki & Brent, 2016), and have the potential to elucidate SA etiology, enabling a more personalized approach to suicide prevention. Finally, given the large number of statistical tests that were completed, future studies examining PRS-ROI associations in larger sample sizes are required to disambiguate potentially spurious associations. Further, while the availability of independent cohorts that include ‘high-risk’ offspring are sparse (de Zwarte et al., 2019), replication of the present findings within a separate case-control study that includes both BD and ‘high-risk’ individuals is required to establish the reliability and generalisability of the presently observed PRS-ROI associations.

## Conclusion

Suicide behaviors remain a significant health burden in BD and are a leading cause of mortality for young people globally. Currently available clinical models of SA have demonstrated poor predictive value, precipitating the search for viable biomarkers that will reliably predict suicidal behaviors and illuminate the ideation-to-action pathway. The identification of biological features and their interactions associated with SA represents important targets for reducing the mortality gap in BD; facilitating future identification of high-risk individuals who may benefit most from prevention strategies (reviewed in Bahji et al., 2021; Katz et al., 2013; Mann et al., 2005; Robinson et al., 2018) and thereby reducing suicide mortality within vulnerable populations. Through the examination of associations between regional brain structure and polygenic risk for SA and risky behavior, the present study provides tentative evidence for the role of the anterior cingulate, parahippocampal and cuneus structures in SA. Future exploration of structural imaging phenotypes that are proximal to suicide events and polygenic risk, informed by larger, multi-trait GWAS, may clarify biomarkers for SA that offer predictive value within a clinical setting.

## Data Availability

The data that support the findings of this study are available in the UNSW data archive ResData at www.dataarchive.unsw.edu.au, under Research Data Management Plan reference number D0237303, or are available on request from the authors.

## Acknowledgements

This work was supported by the Australian National Health and Medical Research Council (NHMRC) Project Grant 1066177 (JMF, RKL, JIN), Program Grant 1037196 (PBM, PRS) and Investigator Grants 1176716 (PRS) and 1177991 (PBM), and philanthropic support from the Lansdowne Foundation, Paul and Jenny Reid, Good Talk and the Keith Pettigrew Family Bequest. DNA extraction was undertaken by Genetic Repositories Australia (GRA; www.neura.edu.au/GRA), a national genetic repository with funding provided by NHMRC Enabling Grant 401184. We thank Jessica Johnson and Doug Ruderfer (Mt Sinai School of Medicine, New York, USA) for assistance with sample preparation and PsychArray genotyping, which was undertaken by the Icahn Institute for Genetics and Genomic Sciences, Mt Sinai School of Medicine, New York, USA. We also acknowledge the contribution of Anna Heath (NeuRA) for the preparation of blood samples, Andrew Frankland (UNSW) for data management and extraction, and Pui Ka Yeung and Lidan Zhang (NeuRA) for completion of structural MRI quality control procedures. JMF gratefully acknowledges the Janette Mary O’Neil Research Fellowship in support of this work. CT is a recipient of a “Ramón y Cajal” fellowship (RYC2018-024106-I) from the Spanish MINECO. This research was undertaken with the assistance of resources from the National Computational Infrastructure (NCI), which is supported by the Australian Government. The National Computational Merit Allocation Scheme (NCMAS) & UNSW High Performance Computing (HPC) Resource Allocation Scheme provided support of em5 project (Neuroscience Research Australia, Neurogenetics; PI A/Prof Janice M Fullerton). We thank Martin Thompson (UNSW) and Andrew Cartwright (NeuRA) for assistance with project setup and data transfer to the Gadi HPC workspace.

## Data Availability Statement

The data that support the findings of this study are available in the UNSW data archive “ResData” at www.dataarchive.unsw.edu.au, under Research Data Management Plan reference number D0237303, or are available on request from the authors.

