## Supplementary material for "Effects of polygenic risk for suicide attempt and risky behavior on brain structure in young people with familial risk of bipolar disorder": Supplmental Figures S1-S2

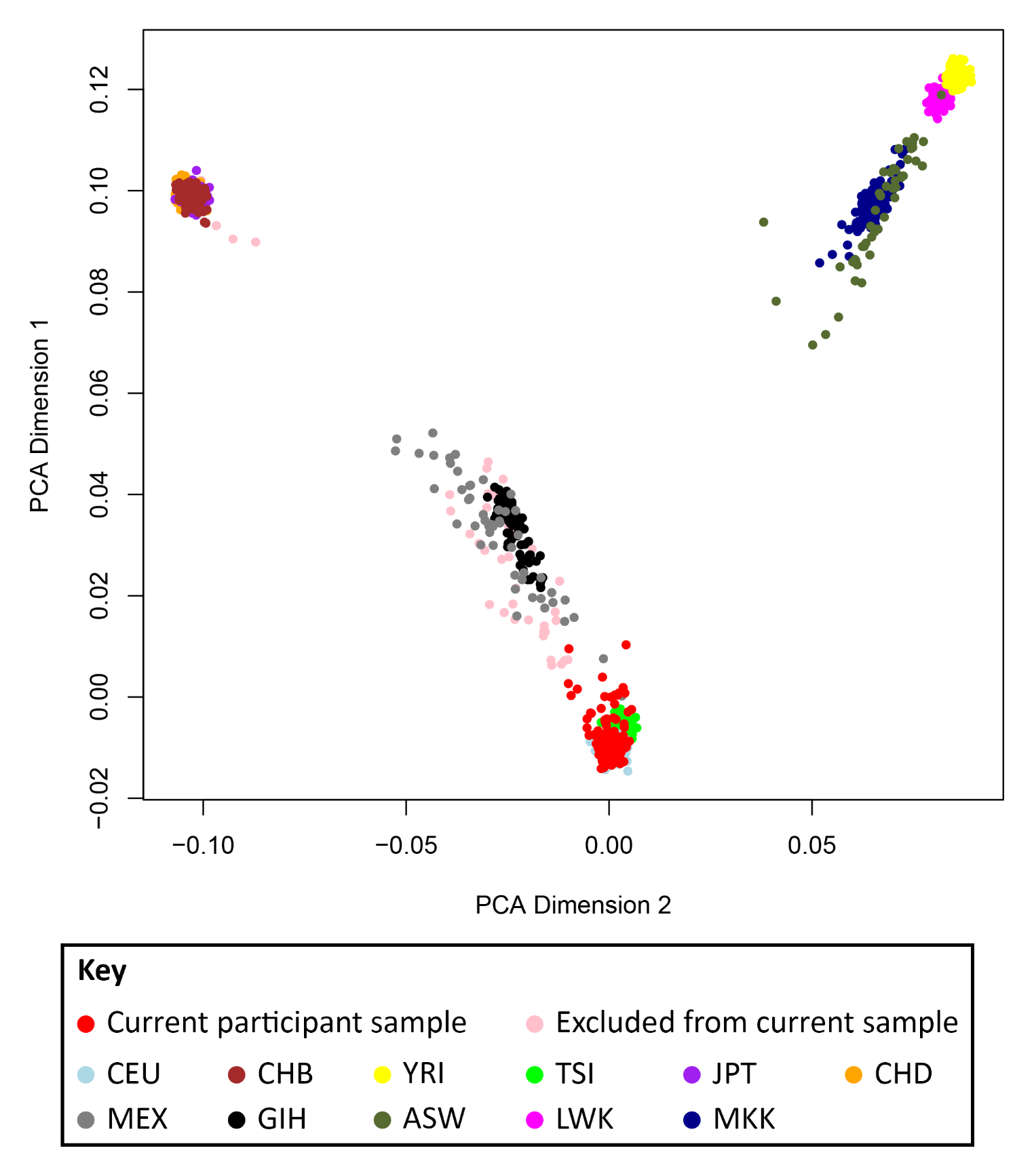

*Figure S1.* Multidimensional scaling plot displaying participant clusters relative to ethnicity principle components 1 and 2. Abbreviation: CEU, Utah residents with northern and western European ancestry; CHB, Han Chinese in Beijing (China); YRI, Yoruba in Ibadan (Nigeria); TSI, Toscani (Italy); JPT, Japanese in Tokyo (Japan); CHD, n=3 Chinese in Denver (Colorado); MEX, Mexican ancestry in Los Angeles (California); GIH, Gujarati Indian in Houston (Texas); ASW, African ancestry in Southwest United States of America; LWK, Luhya in Webuye (Kenya); MKK, Maasai in Kinyawa (Kenya).

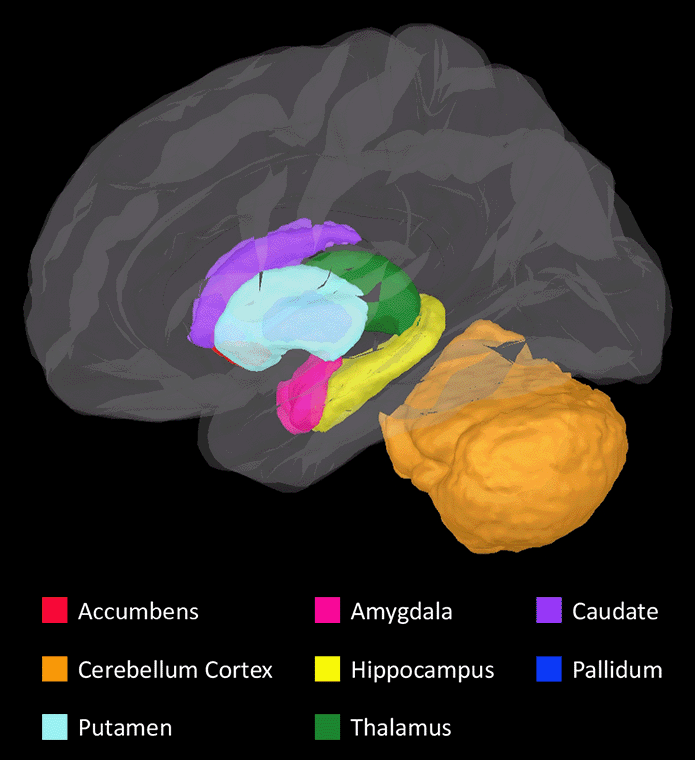

*Figure S2.* Three dimensional horizontally rotating image displaying the location and boundaries of all target subcortical regions-of-interest, selected for analysis in this study on the basis of the literature review of structural neuroimaging studies in suicide attempt. Image was created using Matlab-based program 'Brainstorm' (Mathworks Inc., Massachusetts, USA; Tadel F, Baillet S, Mosher JC, Pantazis D, Leahy RM [2011]. Brainstorm: a user-friendly application for MEG/EEG analysis. Comput Intell Neurosci. 2011:879716. doi: 10.1155/2011/879716).
